# Pre-activity glycemic prediction prioritizes post-meal movement

**DOI:** 10.64898/2026.06.22.26356272

**Authors:** Smadar Shilo, Gal Sapir, Guy Lutsker, Yeela Talmor-Barkan, Anastasia Godneva, Alon Diament, Marcos Matabuena, Eran Segal, Hagai Rossman

## Abstract

Post-meal activity can attenuate glucose excursions; however, the exact magnitude of this effect remains unquantified, and guidance is rarely personalized to the meal occasion. We linked Human Phenotype Project diet logs, continuous glucose monitoring and wearable step counts to test whether glycemic risk estimated before activity occurs can prioritize post-meal movement. An activity-blind PPGR model trained on 391,214 PPGR-valid meals from 9,561 participants generated pre-activity meal scores. Among 55,949 step-linked meals from 1,627 adults without diabetes, higher 0-120-min post-meal steps were associated with lower within-participant PPGR (−53.0 mg/dL*min per 1 s.d. higher log steps; 95% CI, −64.2 to −41.7), with larger adjusted PPGR iAUC contrasts at 1,501-2,500 observed steps (−154.4 mg/dL*min versus 0-50 steps). Associations were stronger among participants with higher glycemic-adiposity burden and after meals with higher predicted PPGR. A held-out pre-activity step-response ranking concentrated larger inverse step-PPGR associations (−79.1 top versus −15.0 mg/dL*min bottom quintile), providing a testable strategy for prediction-guided, post-meal movement prompts.

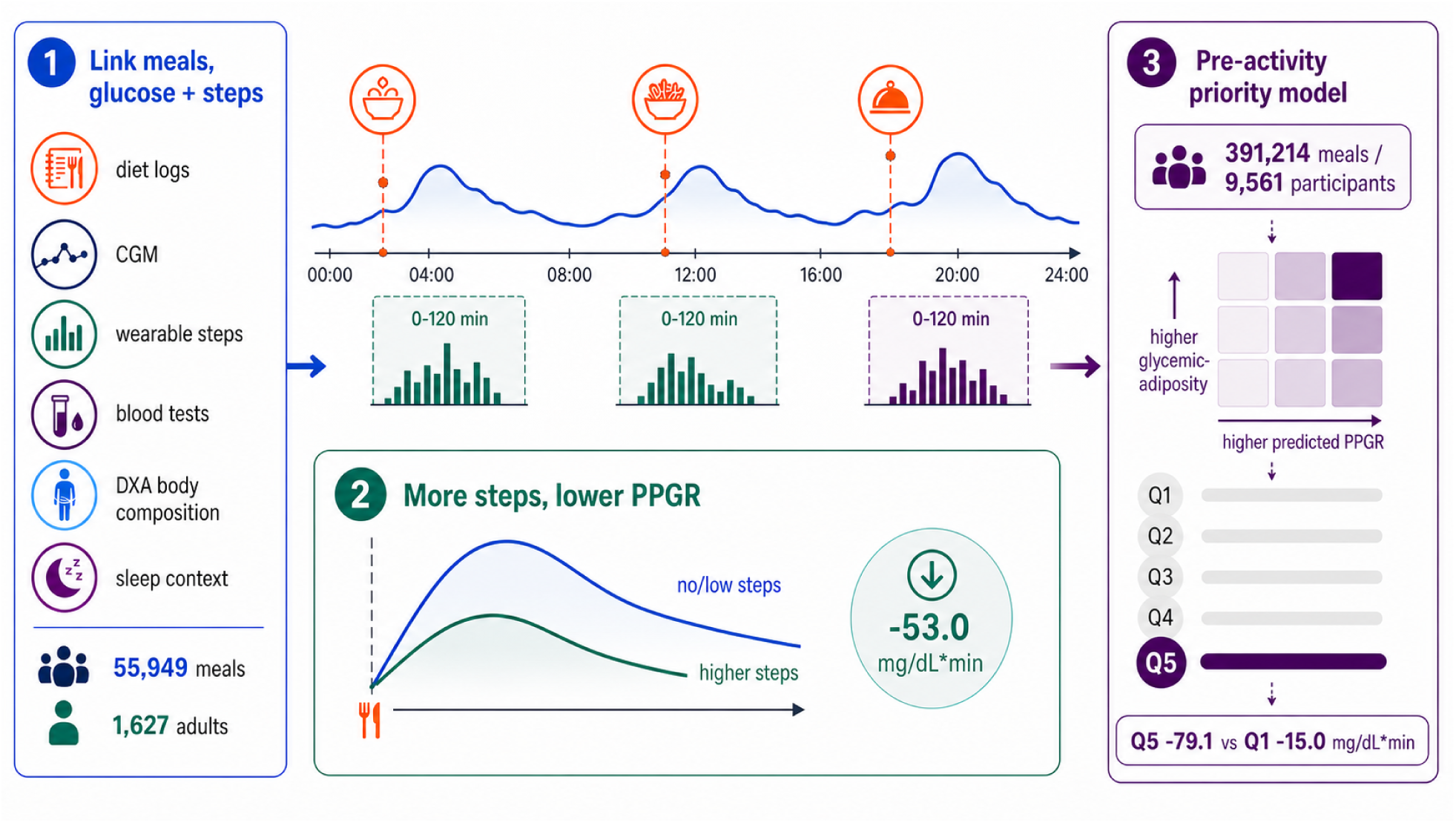

## 1. Introduction

The widespread adoption of continuous glucose monitoring (CGM) has transformed metabolic research by enabling the high-resolution tracking of diverse glycemic features beyond static baseline metrics (Keshet et al. 2023). Meal-related glucose excursions are a dynamic component of everyday glycemic exposure that CGM can resolve more directly than fasting or single-time-point measurements (Zeevi et al. 2015). Randomized controlled trials support the biological premise that walking or other light-to-moderate activity after eating can attenuate postprandial glucose excursions, with stronger evidence for post-meal than pre-meal activity in short-term crossover trials, including randomized studies (Engeroff et al. 2023; Aqeel et al. 2020). Smaller trials using CGM or capillary glucose, including controlled-meal protocols and post-meal walking, show similar directional effects for low-intensity walking, repeated post-meal walking bouts and immediate post-breakfast activity (Manohar et al. 2012; DiPietro et al. 2013; Solomon et al. 2020; Brian et al. 2024).

Whether this experimental pattern is detectable during daily-life meals, naturally chosen step counts and variable daily routines remains uncertain. Yao et al. analyzed 11,333 daily real-life meals from 789 adults without diabetes and used wrist accelerometry to quantify postprandial light and moderate-to-vigorous activity; longer postprandial activity was associated with lower 2-hour postprandial glucose iAUC (Yao et al. 2024). Daily wearable studies also support links between physical activity and CGM-derived glycemic control, although often at a day-level rather than meal-level scale (El Fatouhi et al. 2022). In addition, timing trials do not identify one universally optimal timing strategy after eating: some support immediate activity, whereas others suggest benefit from activity timed closer to the glucose rise or peak (Reynolds and Venn 2018; Hashimoto et al. 2025).

Personalized nutrition studies provide a rationale for targeting post-meal activity to specific eating occasions. CGM, diet, clinical and microbiome features can predict postprandial glycemic responses better than calorie or carbohydrate content alone, and response heterogeneity reflects both meal properties and host physiology (Zeevi et al. 2015; Berry et al. 2020; Mendes-Soares et al. 2019; Wu et al. 2025). However, thus far, these models have mainly been used to guide dietary choices.

The Human Phenotype Project (HPP) provides a high-resolution setting to study the association between meal-related glucose excursions and post-meal activity, as it links large-scale diet logs, CGM, wearable step data and deep phenotyping collected over a two-week monitoring period in the same participants (Shilo et al. 2021; Reicher et al. 2025). Here, we used HPP to test two related questions. First, in 55,949 meals from 1,627 participants with overlapping diet logs, CGM and wearable step counts, we tested whether meals followed by more 0-120 minute steps had lower CGM-derived postprandial glucose response (PPGR) than meals from the same participant followed by fewer steps. This observational, meal-level self-controlled design used participant fixed effects and adjustment for measured meal composition, pre-meal glucose state, meal timing and baseline CGM context, so the primary contrast was within person rather than between more active and less active participants.

Second, we tested whether that association could be prioritized before post-meal movement occurred. We trained an activity-blind predicted-PPGR model in 391,214 HPP meals with valid PPGR outcomes, joined the resulting pre-activity predictions to the step-linked analysis meals, and tested whether the step-PPGR association was stronger in higher predicted-response meals and in participants with higher glycemic-adiposity burden. We then combined meal and participant context into a held-out pre-activity step-response ranking. Robustness, matched-meal, dose-response, timing, CGM-curve-shape and negative-control analyses evaluated the stability and post-meal specificity of the overall and prioritized associations. Full cohort construction, alignment rules, covariate definitions and model equations are detailed in the Online Methods.

## 2. Results

### 2.1. Cohort and meal-level exposure

The primary cohort was drawn from HPP, a prospective deep-phenotyping cohort with longitudinal lifestyle, nutrition, clinical, CGM, sleep and imaging data. In this study, diet logs defined meal timing and composition, Abbott FreeStyle Libre Pro CGM readings sampled every 15 minutes provided pre-meal glucose state and PPGR outcomes, and HealthKit StepCount records quantified post-meal movement. Blood-test, baseline CGM, DXA body-composition and sleep measures provided participant context for cohort description and modifier analyses. The primary analysis included 55,949 estimable meals from 1,627 non-diabetic participants after excluding participants with diabetes diagnoses or glucose-lowering medication use. The cohort was generally middle-aged, while meal-level post-meal movement and macronutrient intake varied substantially within and between individuals. The median meal had 354.7 post-meal steps over 0-120 minutes and contained 36.4 g carbohydrate, supporting a within-participant design rather than unadjusted comparisons between more active and less active people (Table 1). The cohort and exclusion flow is shown in Supplementary Figure 1 and Supplementary Tables 1 and 2. Figure 1 illustrates the linked diet-log, CGM and step streams used for aggregate meal-level analyses.

**Figure 1.**
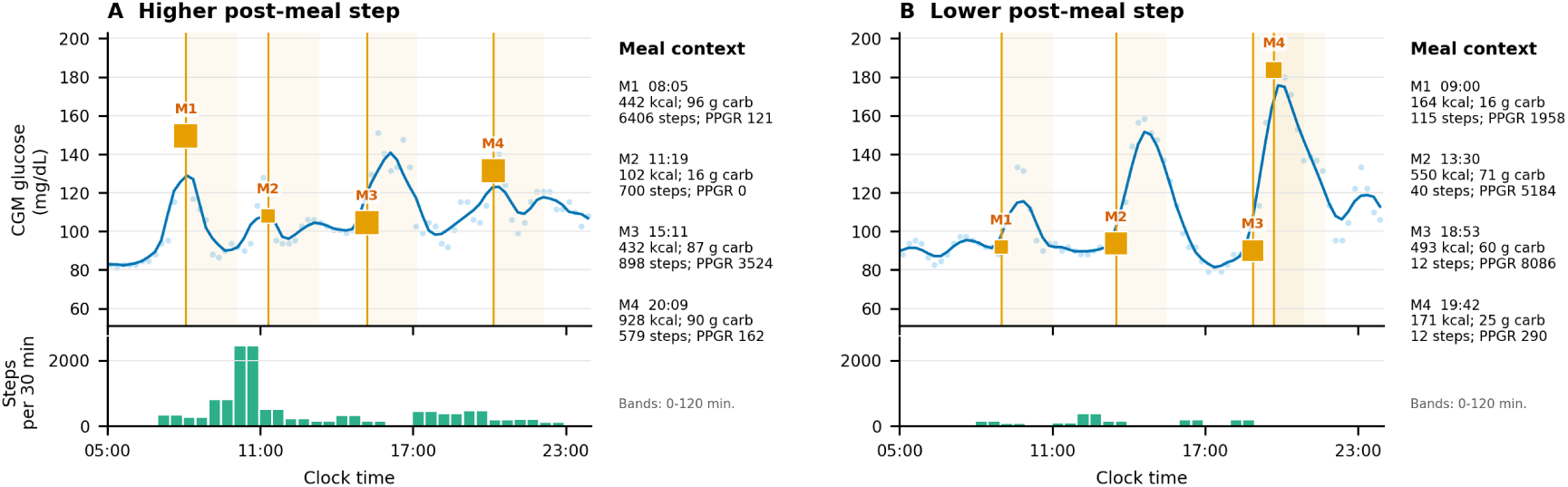
Illustrative participant-days with CGM, meals and steps. Two HPP participant-days show the data streams used to construct meal-level exposures and outcomes. CGM traces are aligned to meal anchors, meal windows mark the 0-120 minute PPGR interval, and wearable step counts are summarized in meal-relative bins. Meal-context tiles provide the measured meal composition, post-meal steps and PPGR iAUC (mg/dL*min) used in cohort-level adjustment and stratification.

**Table 1.** Baseline characteristics of the primary analytic cohort. Participant rows summarize clinical parameters of the study population; meal rows summarize the post-meal activity exposure and diet scale used in the main analysis. Values are n (%), mean (s.d.) or median [IQR], as indicated in the characteristic column. Fasting glucose is the nearest available HPP blood-test glucose measurement to the first analyzed meal. Repeated-meal dietary variability is summarized in Supplementary Table 18 and Supplementary Figure 4.

| characteristic | unit | overall | female | male |
| --- | --- | --- | --- | --- |
| Participants | n | 1627 | 915 | 710 |
| Age | years | 50.1 (9.3) | 49.9 (9.9) | 50.2 (8.5) |
| BMI | kg/m<sup>2</sup> | 25.2 (3.8) | 24.5 (3.9) | 26.1 (3.5) |
| Waist circumference | cm | 85.8 (11.8) | 80.7 (10.4) | 92.3 (10.2) |
| HbA1c | % | 5.3 (0.4) | 5.3 (0.4) | 5.3 (0.4) |
| Fasting glucose | mg/dL | 91.0 (7.9) | 89.9 (7.9) | 92.4 (7.7) |
| Participant baseline CGM mean | mg/dL | 95.9 (10.3) | 94.4 (9.8) | 98.0 (10.6) |
| Sleep AHI | events/hour | 6.5 [3.2, 12.3] | 4.9 [2.4, 9.1] | 9.3 [4.6, 14.5] |
| Meals per participant, median [IQR] | n | 35.0 [24.0, 44.0] | 35.0 [25.0, 43.0] | 36.0 [23.0, 46.0] |
| Participant median post-meal steps, median [IQR] | steps/meal | 340.0 [191.0, 531.0] | 281.0 [151.0, 467.1] | 393.8 [267.0, 595.0] |
| Meals, total | n | 55949 | 31258 | 24590 |
| Post-meal steps 0-120 min, median [IQR] per meal | steps | 354.7 [110.0, 851.0] | 297.0 [81.0, 789.0] | 427.0 [156.0, 926.3] |
| Meal energy, median [IQR] per meal | kcal | 343.7 [198.2, 566.8] | 318.4 [190.0, 510.7] | 382.8 [213.0, 639.6] |
| Carbohydrate, median [IQR] per meal | g | 36.4 [23.7, 59.3] | 34.2 [22.4, 54.6] | 40.4 [25.3, 64.8] |
| Protein, median [IQR] per meal | g | 12.4 [5.1, 25.7] | 11.5 [5.0, 22.7] | 14.0 [5.4, 30.0] |
| Fat, median [IQR] per meal | g | 12.8 [6.1, 24.3] | 11.9 [5.9, 22.2] | 14.0 [6.5, 27.0] |
| Fiber, median [IQR] per meal | g | 4.1 [1.8, 7.7] | 3.9 [1.8, 7.4] | 4.3 [1.9, 8.3] |

### 2.2. Higher post-meal steps were associated with lower PPGR

In the primary within-participant fixed-effect model, higher 0-120 minute post-meal steps were associated with lower 0-120 minute PPGR iAUC (−53.0 mg/dL*min per 1 s.d. higher log-transformed post-meal steps; 95% CI −64.2 to −41.7). This direction was reproduced in the participant-day fixed-effect model (−56.6, 95% CI −70.2 to −43.1), strictly filtered isolated-meal sensitivity (−55.4, 95% CI −70.4 to −40.4) and within-participant matched-meal analysis (−118.7, 95% CI −150.6 to −84.8; Figure 2a). The matched-meal curves showed the same pattern: after matching on measured meal context and pre-meal state, high-step meals had a lower glucose rise through peak and early recovery (Figure 2b). Figure 2b compares related analyses that ask the same question under progressively tighter control: the primary within-participant model compares meals from the same participant; the participant-day fixed-effect model further restricts comparisons to meals from the same person on the same calendar day; the strictly filtered isolated-meal sensitivity applies additional meal-isolation and CGM/diet-quality gates; and the matched-meal analysis compares high-step and low-step meals matched on measured meal context and pre-meal state. Full model definitions are provided in Online Methods.

**Figure 2.**
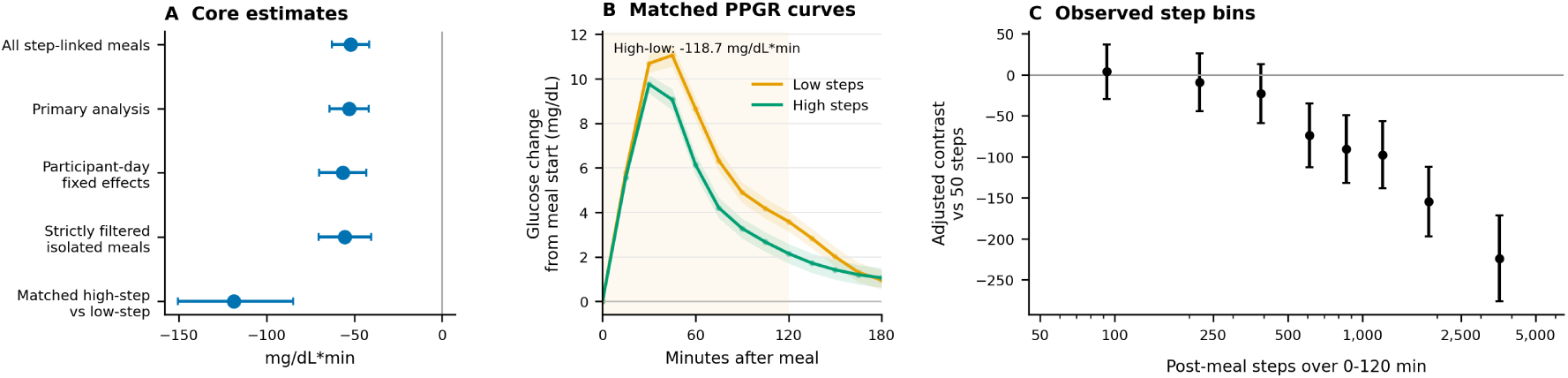
Core step-PPGR estimates, matched glucose curves and granular step-dose contrasts. Panel a summarizes the primary within-participant estimate, participant-day fixed-effect sensitivity, strictly filtered isolated-meal sensitivity and adjusted matched high-step versus low-step meal contrast. Points and horizontal bars are estimates and 95% CIs; negative values indicate lower 0-120 minute PPGR iAUC with higher 0-120 minute post-meal steps, or lower PPGR in matched high-step meals. Panel b shows mean CGM glucose change across all 6,919 within-participant matched meal pairs with CGM-curve support, aligned to the meal anchor from 0 to 180 minutes and zeroed at the meal-start value for display. Low-step meals had 0-250 steps and high-step meals had at least 751 steps in the 0-120 minute post-meal window; shaded bands show approximate 95% CIs for binned means. The high-step curve peaked lower and remained lower through early recovery. Panel c shows adjusted categorical contrasts across observed 0-120 minute step bins (0-50, 51-150, 151-300, 301-500, 501-750, 751-1,000, 1,001-1,500, 1,501-2,500 and 2,500+ steps), plotted at each bin median with vertical pointwise 95% CIs relative to meals followed by 0-50 steps.

### 2.3. Observed step bins showed a graded adjusted PPGR contrast

We next translated the standardized log-step coefficient into observed post-meal step bins, using meals followed by 0-50 steps in 0-120 minutes as the near-sedentary reference. Near the exposure center, 1 s.d. on the log-step scale mapped from about 288 to 1,299 observed post-meal steps. Estimating cumulative meal-level glucose-exposure contrasts (reported in mg/dL*min) over the 0-120 minute PPGR window, meals followed by 51-150 steps differed little from the reference (4.0 mg/dL*min; 95% CI −29.2 to 37.2), and meals followed by 151-300 or 301-500 steps remained close to zero. The contrast became clearly negative at 501-750 steps (−73.6; 95% CI −112.5 to −34.8), 751-1,000 steps (−90.6; 95% CI −132.1 to −49.1), 1,001-1,500 steps (−97.5; 95% CI −138.5 to −56.4), 1,501-2,500 steps (−154.4; 95% CI −197.1 to −111.8) and 2,500+ steps (−223.9; 95% CI −276.4 to −171.4; Figure 2c; Supplementary Table 5).

### 2.4. Stronger associations by participant and meal context

We next tested whether the post-meal step association was related to host metabolic health. The participant-level question was whether people with higher baseline glycemic burden (CGM mean and HbA1c) or adiposity (DXA, BMI, waist circumference) showed a larger association between post-meal steps and lower PPGR. We combined participant baseline CGM mean and HbA1c into a glycemic-burden score, and DXA android/gynoid ratio, percent fat and visceral adipose tissue mass into a DXA adiposity score. We evaluated adiposity in two complementary ways: DXA-derived body composition as the deep-phenotyping measure available in HPP, and BMI and waist circumference as clinically accessible anthropometric modifiers. The DXA adiposity domain was used for the main glycemic-adiposity score, while BMI and waist circumference were reported separately as clinical anthropometric checks. The equal-weight glycemic-adiposity score averaged the glycemic-burden and DXA adiposity domains. It showed a clear gradient: the step association was −32.3 mg/dL*min (95% CI −50.1 to −14.5) in the low tertile, −56.4 mg/dL*min (95% CI −77.0 to −35.8) in the middle tertile and −69.0 mg/dL*min (95% CI −90.3 to −47.8) in the high tertile per 1 s.d. higher log post-meal steps. In the continuous modifier model, the step association was −15.6 mg/dL*min more negative per 1 s.d. higher glycemic-adiposity score (95% CI −27.5 to −3.7). Component checks pointed in the same direction for glycemic burden and DXA percent fat, while BMI and waist circumference were close to zero. Sleep disturbance burden was included as a comparator phenotype and showed no evidence of effect modification. Supporting mixed-model and variance-decomposition analyses were consistent with nonzero participant-level heterogeneity (Supplementary Table 17), so we treated these modifiers as context for prioritization rather than as validated individual treatment effects. A source-level modifier panel also compared these host-physiology estimates with meal-composition and pre-meal-state modifiers, including carbohydrate, protein, fat and pre-meal glucose (Figure 3a-c; Supplementary Tables 9 and 14-17).

**Figure 3.**
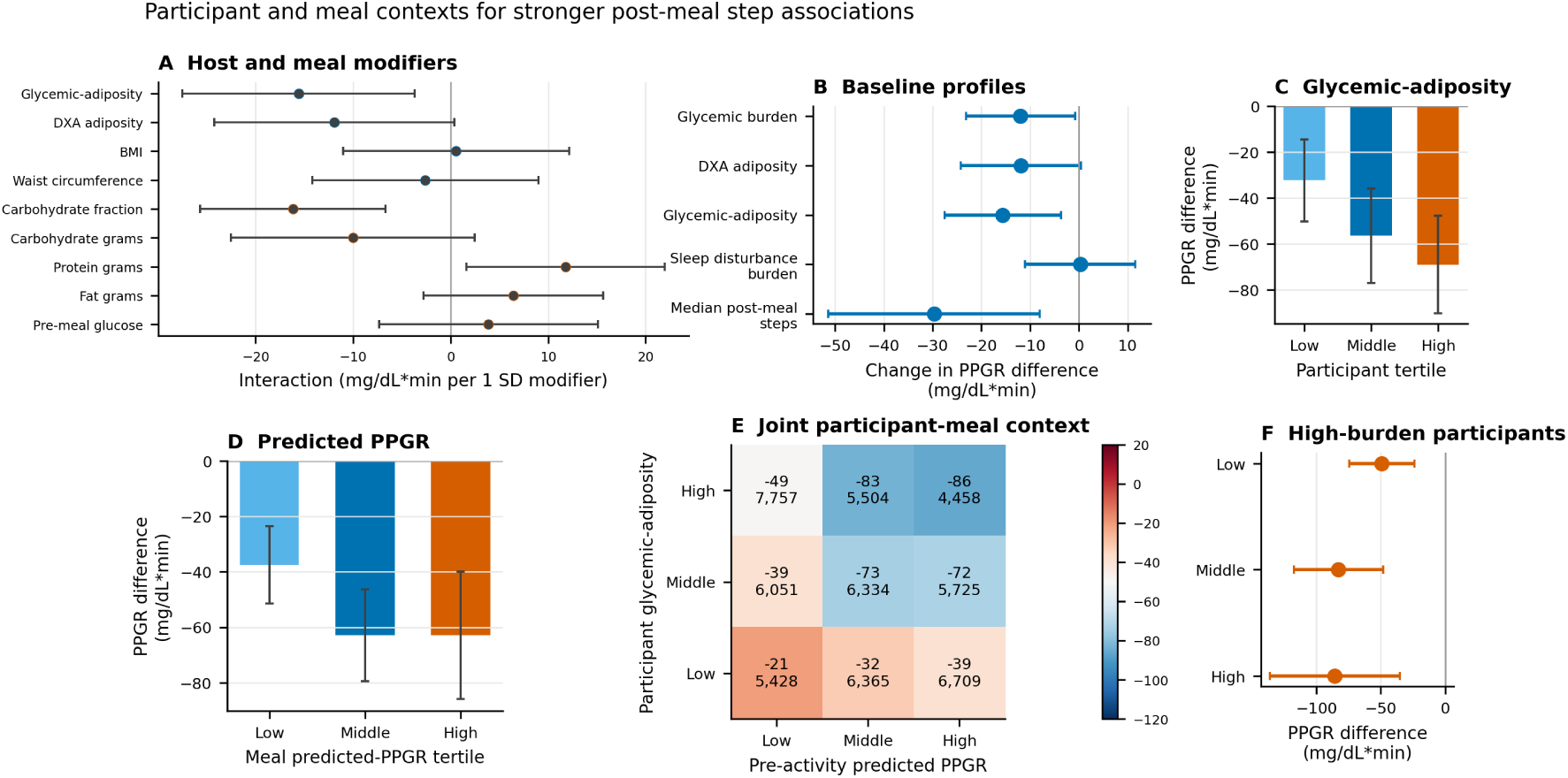
Participant and meal contexts for stronger post-meal step associations. Panel a compares host-physiology and meal-composition/state modifiers of the within-participant association between 0-120 minute post-meal steps and PPGR; more negative interaction estimates indicate stronger lower-PPGR associations at higher modifier values. Panel b shows the main baseline participant profiles as modifiers of the same association. Panel c shows the step association across low, middle and high glycemic-adiposity tertiles. Panel d shows the step association across low, middle and high pre-activity predicted-PPGR meal tertiles. Panel e cross-classifies participant glycemic-adiposity tertile (rows) with pre-activity predicted-PPGR meal tertile (columns); each cell shows the adjusted within-participant association in that subgroup, with meal counts below the estimate. Panel f shows the same step association within high-burden participants across predicted-response meal tertiles. Estimates are in mg/dL*min per 1 s.d. higher log 0-120 minute post-meal steps, or changes in that association per 1 s.d. higher modifier.

We next asked whether specific meal characteristics were associated with the effect of steps on the postprandial glucose response. To test this, we ranked meals by predicted PPGR before any postmeal movement was observed. The ranking score was trained in 391,214 broad HPP PPGR-valid meals from 9,561 participants using meal composition, meal timing, pre-meal glucose state, age, sex, BMI, and nearest HbA1c and fasting glucose blood-test values. These features were included because they are available before post-meal behavior and can rank meals without using activity or observed response fields. In participant-grouped out-of-fold prediction, this pre-activity feature set captured meal-level PPGR variation (R2 = 0.158; mean absolute error = 862.9 mg/dL*min).

We linked the out-of-fold predicted PPGR value to each of the 55,949 meals in the wearable-step analysis set from 1,627 participants. Post-meal steps were more strongly associated with lower PPGR after meals with higher predicted response. In the primary continuous interaction model, the step association became −12.4 mg/dL*min stronger (95% CI −24.5 to −0.3) for each 1 s.d. higher pre-activity predicted PPGR. The same direction was seen with participant-day fixed effects (−9.3, 95% CI −23.2 to 4.6). In tertile terms, the step association strengthened from −37.5 mg/dL*min (95% CI −51.5 to −23.5) in low predicted-response meals to −62.9 mg/dL*min (95% CI −85.8 to −39.9) in high predicted-response meals (Figure 3d; Supplementary Tables 19 and 21).

### 2.5. Combined participant and meal context

We then asked whether the participant-level and meal-level signals overlapped. Participants were first grouped into low, middle and high glycemic-adiposity tertiles, and each meal was independently grouped into low, middle or high pre-activity predicted-PPGR tertiles. We then re-estimated the adjusted within-participant step-PPGR association inside each of the resulting nine cells, so that each cell represents the within-stratum contrast between meals followed by more versus fewer post-meal steps.

The pattern supported the same prioritization logic as the separate modifier analyses. The low-burden/low-response group was close to zero (−20.9 mg/dL*min; 95% CI −43.3 to 1.5), whereas associations were more negative in higher-burden participant strata and in higher predicted-response meal strata. The largest negative estimates were seen among high glycemic-adiposity participants, especially in the middle predicted-response group (−82.9 mg/dL*min; 95% CI −117.6 to −48.1) and high predicted-response group (−85.7 mg/dL*min; 95% CI −136.4 to −35.0; Figure 3e,f; Supplementary Table 22).

### 2.6. A pre-activity step-response ranking concentrated stronger step-PPGR associations

We next asked whether the person-meal context identified above could be turned into a practical ranking: before a meal’s post-meal movement was observed, could we identify occasions where walking was most likely to be associated with a lower glucose excursion? To do this, we combined pre-activity predicted PPGR with participant glycemic-adiposity context into a person-meal step-response score. The score was assigned to held-out meals before post-meal movement was observed and excluded post-meal activity, future activity, post-meal glucose-derived outcomes, next-meal timing and response-informed alignment fields. In held-out meals, the step association became progressively more negative across ranking strata: −15.0 mg/dL*min (95% CI −32.2 to 2.2) in the bottom-ranked quintile and −79.1 mg/dL*min (95% CI −112.6 to −45.5) in the top-ranked quintile per 1 s.d. higher log post-meal steps (Figure 4b).

**Figure 4.**
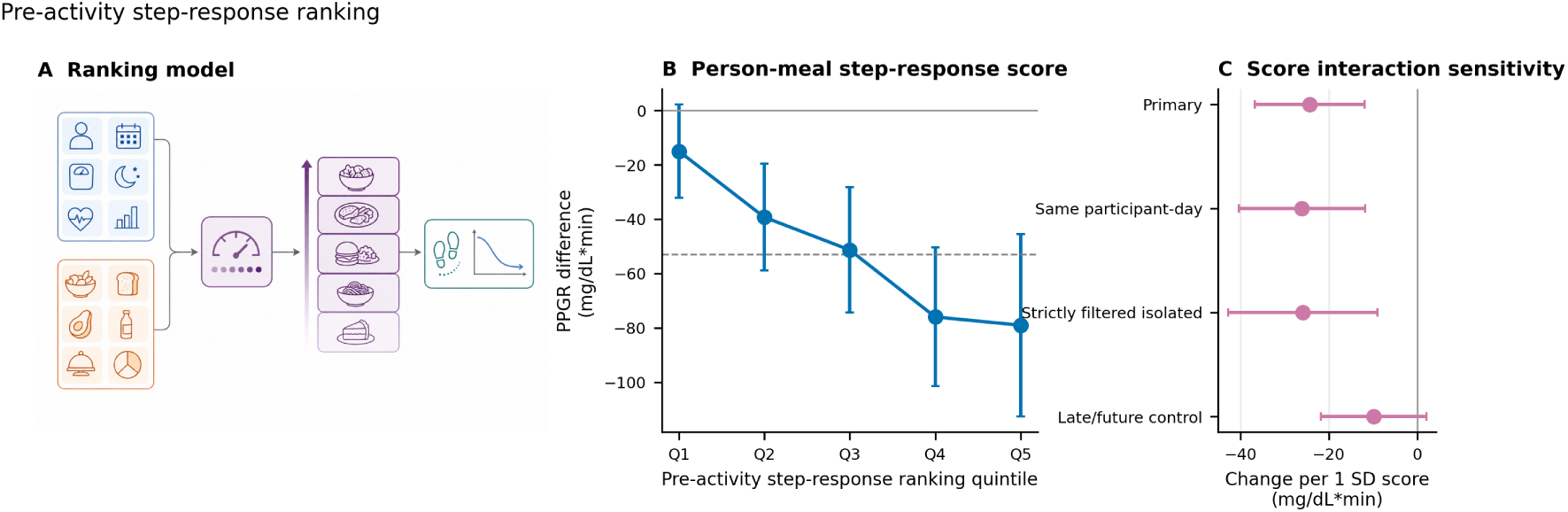
Pre-activity step-response ranking. Panel a illustrates the ranking model: pre-activity participant and meal context are combined before post-meal behavior is observed, meals are ordered into ranking strata, and step-associated PPGR differences are evaluated after ranking. Panel b shows the within-participant association between 0-120 minute post-meal steps and PPGR across held-out quintiles of the selected person-meal step-response score. The score was assigned from pre-activity context only and excluded post-meal movement, future activity, post-meal glucose-derived outcomes and response-informed alignment fields. Points and vertical bars are estimates and 95% CIs in mg/dL*min per 1 s.d. higher log 0-120 minute steps; the dashed line marks the overall primary estimate. Panel c shows continuous score-interaction sensitivity analyses across the primary model, participant-day fixed effects, strictly filtered isolated meals and the late/future activity control. Negative values indicate stronger lower-PPGR associations after meals followed by more post-meal steps.

The continuous score interaction supported the same gradient (−24.3 mg/dL*min per 1 s.d. higher score; 95% CI −36.8 to −11.9), while the late/future activity control crossed zero (−9.9; 95% CI −21.8 to 2.0; Figure 4c). The top-ranked quintile was also negative with participant-day fixed effects. On a practical binary scale, meals with at least 751 post-meal steps had −68.7 mg/dL*min lower adjusted PPGR than meals with at most 250 steps in the bottom-ranked quintile, compared with −186.4 mg/dL*min in the top-ranked quintile. These results provide a practical basis for selecting, in advance, meal occasions for which physical activity is expected to produce a larger reduction in PPGR (Supplementary Tables 23-26).

### 2.7. Sensitivity analyses

We conducted a series of sensitivity analyses to assess the robustness of the primary association. Specifically, we tested whether it persisted under alternative meal-quality criteria, carbohydrate thresholds, step scaling, and model windows, and conducted prespecified control analyses The primary association was preserved in the strictly filtered isolated-meals, participant-day fixed effects, raw-step scale models and carbohydrate-threshold analyses. Meal-context interaction analyses did not indicate that measured meal size alone explained the association. Meal time of day also did not explain the main estimate: removing cyclic meal-hour adjustment changed the primary estimate from −53.0 to −48.4 mg/dL*min per 1 s.d. higher log steps. Clock-time interaction terms suggested some time-of-day variation (joint P = 0.001), with broad strata showing weaker morning and midday estimates and a more negative evening estimate, so we treat meal time as a modifier/sensitivity result rather than as a replacement for the main model. These timing results suggest possible clock-time variation in the step-PPGR association, but they should be interpreted as hypothesis-generating and not as evidence for a specific best time of day for activity recommendations. Calendar date, used as a pre-exposure control for secular drift, was centered near zero. Curve-shape analyses were directionally consistent with the main result: higher post-meal steps were associated with lower early, mid and late post-meal glucose exposure. These supporting results are summarized in Supplementary Tables 8-13 and 28 and Supplementary Figure 3.

## 3. Discussion

This study shows that post-meal movement can be targeted at the meal occasion, before the post-meal behavior has occurred. In 55,949 free-living meals from 1,627 HPP adults without diabetes, meals followed by more 0-120 minute steps had lower adjusted PPGR within the same participant. The association was consistent across participant-day fixed effects, strictly filtered isolated-meal analyses, raw-step scale sensitivity, practical observed-step bins and matched high-step versus low-step meals, and it was not mirrored by negative controls. It was also concentrated in clinically interpretable contexts: higher baseline glycemic-adiposity, higher pre-activity predicted PPGR and a held-out pre-activity step-response ranking all identified person-meal contexts with larger inverse step-PPGR associations. These findings indicate that the effect of post-meal steps on PPGR varies across meal occasions, offering a practical basis for identifying in advance the meals at which physical activity is likely to yield the greatest reduction in PPGR.

These results connect controlled post-meal walking evidence with daily-life meals, variable routines and naturally chosen movement (Engeroff et al. 2023; Brian et al. 2024; Yao et al. 2024). Muscle contraction after eating can increase skeletal-muscle glucose uptake through insulin-independent and GLUT4-related contraction pathways, with related hemodynamic and intracellular signaling mechanisms supporting the biological direction of the observed associations (Richter and Hargreaves 2013). Beyond confirming that post-meal walking is associated with lower glucose exposure, we map this association across tens of thousands of free-living meals and concentrate it into actionable meal-level contexts using diet logs, CGM, wearable steps, and within-person modeling.

The personalization results extend precision nutrition from food selection toward activity timing. Prior PPGR models predict glycemic responses from meal and host features and can outperform population-average nutritional metrics (Zeevi et al. 2015; Berry et al. 2020; Mendes-Soares et al. 2019; Wu et al. 2025). We applied the same modelling principle to a different behavioral decision, shifting the question from what to eat to when post-meal activity is likely to matter most.The predicted-PPGR score ranked meals before post-meal movement was observed and without using post-meal activity, preserving the temporal structure needed for decision support. HPP deep phenotyping supplied a complementary participant axis, with stronger associations among participants with higher CGM/HbA1c glycemic burden and DXA adiposity in a cohort designed for linked CGM, lifestyle and physiologic profiling (Shilo et al. 2021; Reicher et al. 2025).

The modifier pattern adds a practical interpretation to the overall association. Meal composition, pre-meal glucose state and meal timing are established determinants of PPGR, but the observed step association was not explained by measured meal context alone. Instead, higher predicted PPGR and higher glycemic-adiposity burden marked contexts where the inverse step-PPGR association was stronger, consistent with the idea that post-meal movement may matter most when the expected glycemic excursion or baseline metabolic burden is higher. The meal-time sensitivity suggests that clock time may also shape the magnitude of the association, but not enough to account for the primary result. BMI and waist circumference remain clinically accessible checks on this pattern, whereas the DXA-based adiposity score uses the deeper body-composition phenotyping available in HPP.

The pre-activity step-response ranking is the most direct translational bridge from these observational data to an intervention. The score used information available at or before the meal, assigned held-out meals to ranking strata, and then re-estimated the within-person step association within each stratum. Top-ranked meals showed a substantially larger inverse step-PPGR association than bottom-ranked meals, without a corresponding late/future activity signal. This converts heterogeneity into a concrete trial-enrichment rule: prioritize high-ranking meal occasions, deliver a post-meal movement prompt, and estimate prompt uptake and glucose response in a randomized just-in-time design.

The observed-step analyses give that trial design a practical scale. The 751-1,500 and 1,500+ step ranges overlap with short post-meal walking protocols, including 10-minute walks, three 15-minute post-meal bouts and 30-minute brisk postprandial walking interventions (DiPietro et al. 2013; Bellini et al. 2022; Hashimoto et al. 2025). These bins are adjusted observational contrasts rather than prescribed doses, but they show that the standardized model association has a feasible behavioral counterpart in observed step totals. These meal-window bins should not be interpreted as daily-step clinical thresholds. In prospective daily-step literature, higher device-measured step counts are associated with lower all-cause and cardiovascular mortality, with outcome gradients observed below the common 10,000-step target; our 0-120 minute bins therefore provide an occasion-level feasibility scale for a prompt trial rather than a clinical outcome threshold (Paluch et al. 2022; Banach et al. 2023). A prospective study can now test whether prediction-guided prompts after high-priority meals increase post-meal walking and reduce glucose excursions during the first two post-meal hours.

Two design features are central to interpretation. First, the shared 0-120 minute exposure and outcome window captures movement during the post-meal glucose excursion, but it also means that the primary coefficient is a concurrent post-meal movement-glucose association rather than a fully prospective exposure effect. Some steps occurred early enough to plausibly influence the subsequent glucose rise, whereas later steps occurred after part of the glucose response had already unfolded. Matched-meal analyses, timing-distribution summaries and future-activity controls address this issue directly, while randomization remains the appropriate next step for estimating prompt effects.

Future work could model the full post-meal glucose trajectory and activity process jointly, for example with functional-response or time-varying causal frameworks that separate early activity, later activity and evolving glucose dynamics; in the present study, curve-shape, timing-distribution and future-activity controls serve as descriptive checks rather than causal estimators.

Second, measured meal context did not explain the association. Meal composition is a strong determinant of PPGR and a plausible driver of post-meal activity choices (Bellini et al. 2022; Parr et al. 2018; Yao et al. 2024). The primary models adjusted for measured meal composition, and matched high-step and low-step meals remained separated after balancing meal context and pre-meal state. The result is therefore not an unadjusted comparison of different meal types or of generally more active versus less active people.

The study’s strengths are the scale and linkage needed to evaluate this question at meal resolution: 55,949 analyzable wearable-step meals from 1,627 adults, 391,214 broad HPP PPGR-valid meals for pre-activity PPGR prediction, diet-log composition, CGM-derived outcomes, post-meal step-count windows, HPP blood-test, DXA and sleep phenotyping, within-person modeling, matched-meal checks, practical dose translation, pre-activity step-response ranking and negative-control analyses.

The main limitations define the next experimental step. This is within-person observational evidence, so it should not be interpreted as a randomized exercise effect or as a deployable treatment rule. Diet timing and content are self-reported, and self-reported dietary instruments and food-timing recall can carry systematic and random error (Ravelli and Schoeller 2020; Gioia et al. 2022). Step-count event aggregation limits active-minute, bout and sedentary-gap inference; independent 30-minute post-meal step-distribution bins were estimated as coarse overlap-prorated summaries from mostly hourly step-count buckets; and device carry and wear behavior can shape coverage, as shown in smartphone and smartwatch validation studies (Duncan et al. 2018; Amagasa et al. 2019; Hong et al. 2024). Pre-analysis daily step-count summaries rely on observed step-count event support rather than a validated full-day wear-time denominator. Phenotype modifiers and participant-level response phenotypes need reliability validation, and the pre-activity ranking must be tested prospectively before clinical use.

In summary, in this study we show that higher post-meal steps were associated with lower meal-level PPGR within participants, and pre-activity meal and participant context helped identify the meals where this association was largest. These findings suggest that encouraging brief physical activity after eating may be a practical, low-burden strategy to attenuate postprandial glucose excursions, and that incorporating meal and individual context could help target such recommendations to the situations in which they are most likely to be effective.

## 4. Online Methods

### 4.1. Study design, data sources and estimand

We conducted an observational, meal-level analysis within the HPP, a large-scale prospective deep-phenotyping cohort with longitudinal profiling across lifestyle, nutrition, clinical data (e.g. medical diagnosis and medication usage), CGM, sleep and imaging domains (Shilo et al. 2021; Reicher et al. 2025). The analysis unit was a diet-logged meal with overlapping CGM and wearable step-count data. CGM was measured with Abbott FreeStyle Libre Pro sensors worn on the upper arm for up to 14 days, with glucose sampled every 15 minutes. Diet logs, CGM readings and step-count records were summarized relative to the originally logged meal time; no post hoc meal-time correction was applied. The primary exposure was total StepCount in the 0-120 minute interval after the meal anchor, and the primary outcome was baseline-corrected CGM postprandial glucose-response incremental area under the curve (PPGR iAUC) over the same 0-120 minute interval.

The primary estimand was the within-participant meal-to-meal association between post-meal steps and PPGR. Participant fixed effects allowed each participant to serve as their own control, so the primary coefficient compares meals from the same participant that differed in post-meal step exposure after adjustment for measured meal context. This design reduces confounding by stable participant attributes, but it remains susceptible to time-varying factors, including self-selected activity, unmeasured meal details, device carry behavior and residual meal-timing error.

### 4.2. Cohort definition

The primary cohort excluded participants with diabetes diagnoses or glucose-lowering medication use (according to ICD-11 codes 5A11, 5A40 and 5A41 accordingly and medications starting with A10 in ATC code). Meals were included when they had an estimable 0-120 minute PPGR outcome, overlapping wearable step-count data and the covariates required by the corresponding fixed-effect model. No standalone sedentary-participant filter was applied; eligibility required meal-level wearable StepCount support, and low post-meal step counts were retained as part of the modeled exposure distribution. The primary fixed-effect analysis did not impose a minimum number of meals per participant beyond the requirements for estimable within-participant contrasts.

Prespecified sensitivity analyses tested whether the primary estimate depended on meal isolation and stricter CGM/diet-window quality gates. The strictly filtered isolated-meal sensitivity required previous and next meal gaps of at least 120 minutes or unknown, plausible meal size, plausible pre-meal glucose and adequate CGM coverage. Meal-size thresholds for this sensitivity were 50-2,000 kcal and 1-250 g carbohydrate. This sensitivity was designed to test whether the primary association persisted after excluding meals with nearby eating events, extreme reported meal sizes, implausible pre-meal glucose or incomplete CGM support.

### 4.3. Exposure and PPGR outcome construction

Activity was measured from HealthKit StepCount records, a step-count quantity type recorded by compatible devices and apps (StepCount documentation). Published validation work supports the iPhone Health application as a step-count source under laboratory and daily-life conditions, while showing that real-world accuracy can depend on carry behavior and context (Duncan et al. 2018). StepCount was retained as the activity exposure for interpretability, coverage and direct relevance to post-meal walking.

For continuous models, 0-120 minute steps were transformed as l*o*g(1 + s*t*eps) and standardized within the analysis dataset. Practical observed-step bins, raw per-1,000-step sensitivity models and dose-response curves translated the association back to the observed step scale. The primary PPGR iAUC was computed from CGM readings over 0-120 minutes after the meal anchor after subtracting the pre-meal baseline glucose. The pre-meal baseline glucose state was also used as an adjustment covariate.

### 4.4. Primary statistical analysis

The primary fixed-effect model estimated:

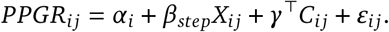

Here, *i* indexes participants, *j* indexes meals, PPG*R_i_*_*j*_ is 0-120 minute PPGR iAUC, *α_i_* is the participant fixed effect, *X_i_*_*j*_ is standardized l*o*g(1+s*t*eps) StepCount from 0-120 minutes after the meal anchor, and *C_i_*_*j*_ is the measured meal-level covariate vector, and *ε_i_*_*j*_ is the residual error term. Covariates included meal composition and size (energy, carbohydrate, protein, fat, fiber and number of foods), pre-meal baseline glucose, meal timing (hour and weekday sine/cosine terms), previous-meal gap and baseline CGM context when available. Numeric covariates were retained when sufficiently observed and variable in the analysis dataset.

The coefficient *β*_s*t*ep_ is the adjusted within-participant difference in PPGR iAUC associated with a 1 s.d. higher log-transformed post-meal step exposure. Negative estimates indicate lower postprandial glucose exposure after meals followed by more steps. Sensitivity models changed the comparison set rather than the scientific estimand. Participant-day fixed effects restricted the comparison to meals eaten by the same participant on the same calendar day. Strictly filtered isolated-meal models applied the additional temporal-isolation and physiologic-plausibility gates described above. Negative-control analyses used late/future activity windows or pre-exposure outcomes outside the immediate post-meal movement contrast.

Uncertainty estimates preserved participant-level dependence. Core fixed-effect estimates used participant-level resampling or participant-clustered robust inference as appropriate for the source model. Follow-up interaction and stratified models used fixed-effect ordinary least squares with robust standard errors clustered by the relevant fixed-effect group. Confidence intervals are reported as 95% intervals throughout.

Observed step-bin analyses translated the primary log-step association back to practical post-meal step totals, comparing each category with a 0-50 step reference bin. A supplemental natural cubic spline of l*o*g(1 + s*t*eps) estimated contrasts relative to 50 post-meal steps in the same participant fixed-effect framework. Detailed dose-response tables and spline settings are reported in the Supplementary Information.

### 4.5. Matched high-step versus low-step meal analysis

The matched-meal analysis compared glucose trajectories for high-step and low-step meals from the same participant after matching on meal composition, pre-meal glucose state, prior meal gap and meal timing. Candidate meals were classified as low-step when they had 0-250 steps and high-step when they had at least 751 steps in the 0-120 minute post-meal window. High-step and low-step meals were paired within participant using measured meal composition, pre-meal glucose state, prior meal gap and meal timing variables. The matched contrast was adjusted for residual pair differences and bootstrapped by participant. Matched CGM curves were reconstructed from meal-aligned CGM readings and summarized as baseline-subtracted binned means across all matched pairs with curve support. Balance diagnostics, matching variables and pre-meal curve checks are reported in Supplementary Note 3.

### 4.6. Participant deep-phenotyping modifiers

Participant-level modifiers were defined before meal-level modeling and then attached to every eligible meal from that participant. Glycemic burden averaged participant-standardized baseline CGM mean and HbA1c when available. DXA adiposity averaged participant-standardized android/gynoid ratio, percent fat and visceral adipose tissue mass when at least two components were available. BMI and waist circumference were retained as clinically available anthropometric component checks but were not included in the DXA adiposity score, which was intended to represent the body-composition imaging domain. The glycemic-adiposity score was the equal-weight average of glycemic burden and DXA adiposity. Sleep disturbance burden, a comparator domain, averaged log-transformed apnea-hypopnea index, lower sleep efficiency, number of wakes and lower minimum oxygen saturation.

The modifier model estimated whether the within-participant meal-to-meal association between postmeal steps and PPGR differed according to baseline phenotype score:

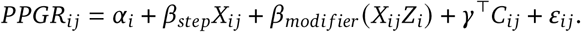

Here, *Z_i_* is the standardized participant-level phenotype score. Because *Z_i_* is constant within participant, its main effect is absorbed by the participant fixed effect. The estimand is therefore *β*_m*o*di*fier*_, the change in the post-meal step slope per 1 s.d. higher phenotype score. A negative *β*_m*o*di*fier*_ means that the inverse within-participant association between post-meal steps and PPGR is stronger among participants with higher baseline phenotype burden. The glycemic-adiposity score was also tested with participant-day fixed effects and strictly filtered isolated meals.

For interpretability, the glycemic-adiposity score was divided into participant-level tertiles and the post-meal step association was estimated separately within each tertile using the same participant fixed-effect framework. These strata describe the gradient of association; the interaction model supplies the formal test of effect modification.

Baseline activity was evaluated as a supplemental participant modifier using StepCount records from the 30 calendar days before each participant’s first primary-analysis meal. Intervals crossing local midnight were allocated to participant-days, and the primary summary was standardized log median daily steps among participants with at least seven observed StepCount days. Sensitivities repeated the analysis with 14-day and 7-day windows and with observed-event-hour filters. Because observed StepCount-event hours are not a validated wear-time denominator, these analyses were treated as supplemental activity-context checks.

### 4.7. Pre-activity predicted PPGR modifier

Pre-activity predicted PPGR was estimated with participant-grouped out-of-fold ridge models in a broad HPP PPGR-valid meal cohort, then joined back to the wearable-step analysis dataset. The prediction source was the non-diabetic HPP meal data with valid PPGR outcomes, anchored at the originally logged meal time and not restricted by post-meal activity availability. The selected model used meal composition (energy, carbohydrate, protein, fat, fiber, number of foods and carbohydrate fraction), meal timing (previous-meal gap, hour and weekday), pre-meal CGM state (baseline glucose), basic participant phenotype (age, sex and BMI), and nearest available glycemic blood-test values (HbA1c and fasting glucose). Numeric features were imputed, standardized and fit with ridge regression.

Activity, active energy, distance, heart rate, other device fields, future activity windows, response-derived glucose outcomes, response-informed alignment fields, omics, genetics, DXA/body composition, habitual CGM summaries and next-meal timing were excluded from the primary predicted-PPGR score. Prediction folds were grouped by participant using ten deterministic folds, so all meals from a participant were held out together when generating that participant’s out-of-fold predictions. The score was therefore used as a pre-activity stratification variable rather than as a mediator of observed post-meal behavior.

Out-of-fold predictions from the selected model were joined to the wearable-step analysis table by exact participant identifier and originally logged meal time. All 55,949 meals in the wearable-step analysis had a matching out-of-fold prediction after join-status and target-agreement checks. The selected predictions were standardized within the joined activity dataset before interaction and tertile analyses. The main interaction model was:

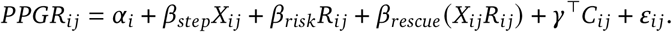

Here, *R_ij_* is the standardized pre-activity predicted PPGR. The estimand *β*_resc*u*e_ is the change in the post-meal step association per 1 s.d. higher predicted meal PPGR. A negative value means that the inverse within-participant association between post-meal steps and PPGR is stronger after meals predicted to have higher PPGR. Sensitivity models used participant-day fixed effects, strictly filtered isolated meals and a late/future activity negative-control exposure.

### 4.8. Joint participant and meal context

The joint context analysis combined participant glycemic-adiposity tertile with pre-activity predicted-PPGR tertile. Tertiles were assigned at the participant level for glycemic-adiposity and at the meal level for predicted meal PPGR. Within each 3 x 3 cell, we estimated the participant fixed-effect association between standardized log post-meal steps and 0-120 minute PPGR iAUC. A continuous three-way model tested the *X_ij_* × *Z_i_* × *R_i_*_*j*_ interaction, with lower-order step-by-predicted-response terms and the predicted-response main effect included; the participant-level glycemic-adiposity main effect was absorbed by participant fixed effects.

### 4.9. Pre-activity step-response ranking

After estimating the overall association between post-meal steps and PPGR, we asked whether this association could be prioritized before post-meal movement occurred. The practical question was not simply which meals were predicted to have higher PPGR. It was: among meals eaten by the same person, can pre-activity information identify the occasions where more-than-usual post-meal steps are most strongly associated with lower-than-usual PPGR?

The goal was to turn this observed heterogeneity into a meal-level ranking for future prompt testing. A high-ranking meal should be interpreted as a meal occasion where post-meal movement appeared more strongly linked to lower PPGR in the observational data, using only information available before movement occurred.

Candidate predictors were restricted to pre-activity information: pre-activity predicted PPGR, meal composition and size, meal timing, previous-meal gap, pre-meal glucose level, age, sex, BMI, nearest HbA1c, nearest fasting glucose and HPP glycemic-adiposity scores. Features derived from post-meal behavior, post-meal glucose, or future events were excluded. The main readout was a person-meal step-response score combining predicted PPGR and glycemic-adiposity context; fully modeled ridge and predicted-response-only scores were evaluated as sensitivities.

The modeling target for the ridge score was the within-participant step-PPGR slope, not PPGR itself.

This distinction matters because a model that only predicts high PPGR would mainly identify large glucose excursions. That is useful for risk stratification, but it does not directly answer where postmeal movement is most strongly associated with lower glucose exposure. The step-response ranking therefore asks a slope question: does the step-PPGR relationship become more negative in some pre-activity contexts than in others?

We used an R-learner-style residualization strategy to make that question explicit. First, within each training fold, we removed each participant’s average PPGR level and average post-meal step tendency:

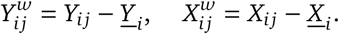

Here, 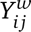 is the meal’s PPGR deviation from that participant’s average PPGR, and 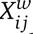 is the meal’s post-meal step deviation from that participant’s average post-meal steps. This residualization keeps the comparison within person. A participant who usually walks more after meals is therefore not automatically assigned a higher response score; the model instead asks whether, for that participant, meals with more-than-usual steps tended to have lower-than-usual PPGR.

After residualization, the model learned which pre-activity features marked a stronger residual step-PPGR slope. The ratio 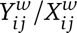 provides a meal-level estimate of that slope, but it is unstable when within-participant step contrast is small. We therefore trained a ridge model on this pseudo-outcome only among meals with sufficient step contrast and weighted meals by 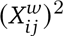, so meals with more informative step differences contributed more to the fitted slope. In this formulation, the model estimates a slope function *τ*(*Z_ij_*), where *Z_ij_* denotes the pre-activity feature vector.

This construction follows the motivation of Robinson residualization and the R-learner objective: remove predictable baseline outcome and exposure components first, then learn how the remaining exposure-outcome slope varies with covariates. In this study, that logic was used to rank observational step-PPGR associations before post-meal movement was observed (Robinson 1988; Nie and Wager 2021; Chernozhukov et al. 2018).

Participant-grouped cross-fitting kept all meals from a participant together when assigning held-out scores. Scores were multiplied by −1 and standardized so that higher values indicated a more negative expected step-PPGR slope.

Evaluation re-estimated the fixed-effect step-PPGR association within held-out score quintiles rather than interpreting the score directly. Sensitivities used participant-day fixed effects, strictly filtered isolated meals, a late/future activity negative-control exposure and high-versus-low step contrasts.

### 4.10. Timing readouts

Timing summaries decomposed available StepCount records into meal-relative 0-30, 30-60, 60-90 and 90-120 minute windows using overlap-prorated raw events, then scaled the four bins to each meal’s tracked 0-120 minute aggregate step total. Meal time-of-day sensitivity compared the primary cyclic meal-hour-adjusted model with an otherwise matched model excluding hour sine/cosine terms, fitted paired step-by-hour sine/cosine interaction terms and repeated the step model within morning, midday, evening and late-night logged-meal strata.

### 4.11. Ethics

HPP participants sign informed consent on arrival at the research site. Identifying details are removed before computational analysis. The cohort is conducted according to the Declaration of Helsinki and was approved by the Weizmann Institute of Science IRB, approval number 2392-4.

## Data availability

Privacy-reviewed aggregate source data underlying the figures, tables and main manuscript claims are provided with the analysis repository where permitted by privacy review. Individual-level Human Phenotype Project diet-log, CGM, wearable step-count and deep-phenotyping data are not publicly distributed because of participant privacy and ethical restrictions set by the study IRB. Bona fide researchers from universities and other research institutions can request access to HPP data through the Human Phenotype Project data access portal at https://humanphenotypeproject.org/data-access or by contacting. Approved access is provided for non-commercial research in a secure cloud-based environment after execution of the required data-use agreement.

## Code availability

Analysis code and privacy-reviewed aggregate outputs needed to reproduce the manuscript figures and tables are maintained at https://github.com/hrossman/pre-activity-glycemic-prediction-prioritizes-post-meal-movement.

## Acknowledgements

We thank the Human Phenotype Project participants and the Human Phenotype Project study team for building and maintaining the cohort infrastructure that made this analysis possible. E.S. is supported by the Crown Human Genome Center; the Larson Charitable Foundation New Scientist Fund; the Else Kroener Fresenius Foundation; the White Rose International Foundation; the Ben B. and Joyce E. Eisenberg Foundation; the Nissenbaum Family; Marcos Pinheiro de Andrade and Vanessa Buchheim; Lady Michelle Michels; Aliza Moussaieff; grants funded by the Minerva Foundation, with funding from the Federal German Ministry for Education and Research; the European Research Council; and the Israel Science Foundation. The funders had no role in study design, data collection and analysis, decision to publish or preparation of the manuscript.

## Author contributions

S.S. and H.R. conceived the study, designed the analyses, developed the methodology, performed the investigation, interpreted the results, wrote the manuscript and supervised the project; G.S. developed the methodology, reviewed the analyses, interpreted the results and critically revised the manuscript; G.L., Y.T.B., A.G., A.D., M.M. and E.S. reviewed the analyses, interpreted the results and critically revised the manuscript. All authors approved the final manuscript.

## References

1. Keshet A, et al. CGMap: Characterizing continuous glucose monitor data in thousands of non-diabetic individuals. Cell Metabolism. 2023. 10.1016/j.cmet.2023.04.002

2. Engeroff T, et al. After dinner rest a while, after supper walk a mile? A systematic review with meta-analysis on the acute postprandial glycemic response to exercise before and after meal ingestion in healthy subjects and patients with impaired glucose tolerance. Sports Medicine. 2023. 10.1007/s40279-022-01808-7

3. Aqeel M, et al. The effect of timing of exercise and eating on postprandial response in adults: a systematic review. Nutrients. 2020. 10.3390/nu12010221

4. Manohar C, et al. The effect of walking on postprandial glycemic excursion in patients with type 1 diabetes and healthy people. Diabetes Care. 2012. 10.2337/dc11-2381

5. DiPietro L, et al. Three 15-min bouts of moderate postmeal walking significantly improves 24-h glycemic control in older people at risk for impaired glucose tolerance. Diabetes Care. 2013. 10.2337/dc13-0084

6. Solomon TPJ, et al. Immediate post-breakfast physical activity improves interstitial postprandial glycemia: a comparison of different activity-meal timings. Pflugers Archiv - European Journal of Physiology. 2020. 10.1007/s00424-019-02300-4

7. Brian MS, et al. Post-meal exercise under ecological conditions improves post-prandial glucose levels but not 24-hour glucose control. Journal of Sports Sciences. 2024;42:728–736. 10.1080/02640414.2024.2363688

8. Reynolds AN, Venn BJ. The timing of activity after eating affects the glycaemic response of healthy adults: a randomised controlled trial. Nutrients. 2018. 10.3390/nu10111743

9. Hashimoto T, et al. Positive impact of a 10-min walk immediately after glucose intake on postprandial glucose levels. Scientific Reports. 2025. 10.1038/s41598-025-07312-y

10. Dunstan DW, et al. Breaking up prolonged sitting reduces postprandial glucose and insulin responses. Diabetes Care. 2012. 10.2337/dc11-1931

11. Loh R, et al. Effects of interrupting prolonged sitting with physical activity breaks on blood glucose, insulin and triacylglycerol measures: a systematic review and meta-analysis. Sports Medicine. 2020. 10.1007/s40279-019-01183-w

12. Quan M, et al. Effects of interrupting prolonged sitting on postprandial glycemia and insulin responses: a network meta-analysis. Journal of Sport and Health Science. 2021. 10.1016/j.jshs.2020.12.006

13. Zeevi D, et al. Personalized nutrition by prediction of glycemic responses. Cell. 2015. 10.1016/j.cell.2015.11.001

14. Berry SE, et al. Human postprandial responses to food and potential for precision nutrition. Nature Medicine. 2020;26:964–973. 10.1038/s41591-020-0934-0

15. Mendes-Soares H, et al. Model of personalized postprandial glycemic response to food developed for an Israeli cohort predicts responses in Midwestern American individuals. American Journal of Clinical Nutrition. 2019. 10.1093/ajcn/nqz028

16. Wu Y, et al. Individual variations in glycemic responses to carbohydrates and underlying metabolic physiology. Nature Medicine. 2025. 10.1038/s41591-025-03719-2

17. Yao J, et al. Diet, physical activity, and sleep in relation to postprandial glucose responses under free-living conditions: an intensive longitudinal observational study. International Journal of Behavioral Nutrition and Physical Activity. 2024. 10.1186/s12966-024-01693-5

18. El Fatouhi D, et al. Associations between device-measured physical activity and glycemic control and variability indices under free-living conditions. Diabetes Technology & Therapeutics. 2022. 10.1089/dia.2021.0294

19. Shilo S, et al. 10K: a large-scale prospective longitudinal study in Israel. European Journal of Epidemiology. 2021. 10.1007/s10654-021-00753-5

20. Reicher L, et al. Deep phenotyping of health-disease continuum in the Human Phenotype Project. Nature Medicine. 2025. 10.1038/s41591-025-03790-9

21. Richter EA, Hargreaves M. Exercise, GLUT4, and skeletal muscle glucose uptake. Physiological Reviews. 2013;93:993–1017. 10.1152/physrev.00038.2012[1]

22. Bellini A, et al. The effects of postprandial walking on the glucose response after meals with different characteristics. Nutrients. 2022. 10.3390/nu14051080

23. Paluch AE, et al. Daily steps and all-cause mortality: a meta-analysis of 15 international cohorts. Lancet Public Health. 2022. 10.1016/S2468-2667(21)00302-9[2]

24. Banach M, et al. The association between daily step count and all-cause and cardiovascular mortality: a meta-analysis. European Journal of Preventive Cardiology. 2023;30:1975–1985. 10.1093/eurjpc/zwad229[3]

25. Parr EB, et al. Effects of providing high-fat versus high-carbohydrate meals on daily and postprandial physical activity and glucose patterns: a randomised controlled trial. Nutrients. 2018. 10.3390/nu10050557

26. Ravelli MN, Schoeller DA. Traditional self-reported dietary instruments are prone to inaccuracies and new approaches are needed. Frontiers in Nutrition. 2020. 10.3389/fnut.2020.00090

27. Gioia SC, et al. How accurately can we recall the timing of food intake? A comparison of food times from recall-based survey questions and daily food records. Current Developments in Nutrition. 2022. 10.1093/cdn/nzac002

28. Duncan MJ, Wunderlich K, Zhao Y, Faulkner G. Walk this way: validity evidence of iPhone health application step count in laboratory and free-living conditions. Journal of Sports Sciences. 2018;36:1695–1704. 10.1080/02640414.2017.1409855

29. Amagasa S, et al. How well iPhones measure steps in free-living conditions: cross-sectional validation study. JMIR mHealth and uHealth. 2019. 10.2196/10418

30. Spartano NL, et al. Agreement Between Apple Watch and Actical Step Counts in a Community Setting: Cross-Sectional Investigation From the Framingham Heart Study. JMIR Biomedical Engineering. 2024;9:e54631. 10.2196/54631[4]

31. Apple Inc. HKQuantityTypeIdentifier.stepCount. Apple Developer Documentation. https://developer.apple.com/documentation/healthkit/hkquantitytypeidentifier/stepcount

32. Robinson PM. Root-N-consistent semiparametric regression. Econometrica. 1988;56:931-954. 10.2307/1912705

33. Nie X, Wager S. Quasi-oracle estimation of heterogeneous treatment effects. Biometrika. 2021;108:299–319. 10.1093/biomet/asaa076

34. Chernozhukov V, Chetverikov D, Demirer M, Duflo E, Hansen C, Newey W, Robins J. Double/debiased machine learning for treatment and structural parameters. The Econometrics Journal. 2018;21:C1–C68. 10.1111/ectj.12097

