## supplementary for "Pre-activity glycemic prediction prioritizes post-meal movement"

### Supplementary Information

This Supplementary Information provides cohort construction, robustness, sensitivity, heterogeneity and prediction-model results supporting the post-meal physical activity and PPGR manuscript.

#### Supplementary Note 1: Cohort Construction and Analysis Set

The analysis set was constructed from logged meals with CGM and wearable step-count support. Participant exclusions removed diabetes diagnoses and glucose-lowering medication use, and meal timing was anchored to the originally logged meal time.

**Supplementary Table 1** | Cohort construction from logged meals with step-count support to the primary analysis and strictly filtered sensitivity cohort.

| stage | meals | participants | median meals per participant |
|---|---|---|---|
| Logged meals with step-count support | 92,625 | 1,883 | 49.0 |
| Meals with valid PPGR outcome | 92,260 | 1,883 | 49.0 |
| Meals with plausible calories and carbohydrates | 91,597 | 1,883 | 49.0 |
| After glucose-lowering medication exclusion | 85,422 | 1,758 | 49.0 |
| After diabetes diagnosis exclusion | 55,949 | 1,627 | 35.0 |
| Strictly filtered isolated-meal sensitivity cohort | 30,108 | 1,614 | 18.0 |

**Supplementary Table 2** | Participant exclusions and analysis-set coverage used to define the step-linked cohort.

| category | measure | value | unit |
|---|---|---|---|
| Cohort size | participants with logged meals | 1,883 | participants |
| Cohort size | meals with valid PPGR outcome | 92,260 | meals |
| Cohort size | meals with wearable-step support | 64,869 | meals |
| Cohort size | participants with wearable-step support | 1,875 | participants |
| Primary analysis | meals | 55,949 | meals |
| Primary analysis | participants | 1,627 | participants |
| Strictly filtered sensitivity | meals | 30,108 | meals |
| Primary analysis | median 0-120 min post-meal steps | 354.8 | steps |
| Primary analysis | median carbohydrate per meal | 36.4 | g |
| Primary analysis | median 0-120 min PPGR iAUC | 904.5 | mg/dL*min |
| Cohort size | logged meals entering meal-level checks | 92,625 | meals |
| Participant exclusions | participants evaluated for exclusion | 29,820 | participants |
| Participant exclusions | participants with glucose-lowering medication use | 1,126 | participants |
| Participant exclusions | participants with diabetes diagnosis | 2,408 | participants |
| Participant exclusions | participants with any diabetes-related exclusion | 2,904 | participants |
| Participant exclusions | medication fields contributing to glucose-lowering medication screen | Metformin Teva; Januet; Januvia; Glucomin | fields |
| Participant exclusions | diabetes diagnosis rule | ICD-like medical-condition code begins with 5A10-5A14 or 5A40 | rule |
| Participant exclusions | glucose-lowering medication rule | ATC code begins with A10, or medication-name keyword indicates glucose-lowering treatment | rule |

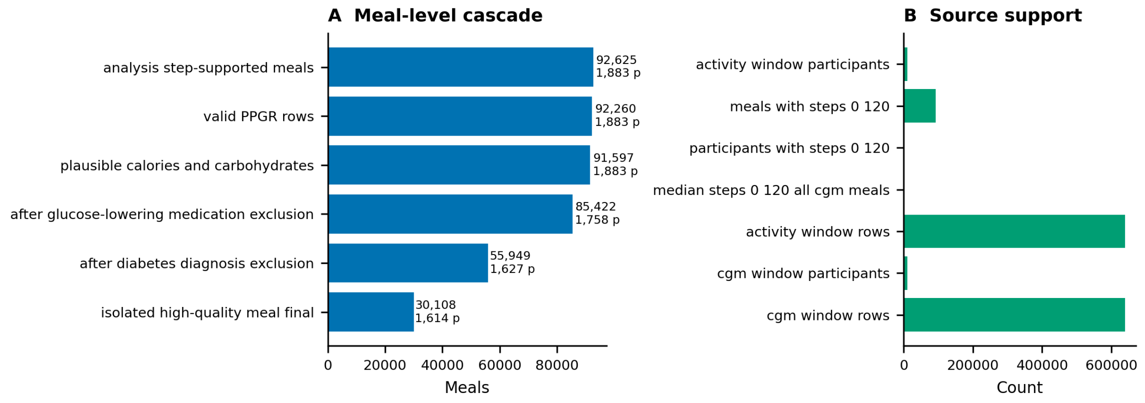


**Supplementary Figure 1** | Cohort cascade and data coverage. Bars summarize aggregate meal inclusion steps and available source support for the step-linked analysis set.

#### Supplementary Note 2: Core Robustness and Exposure Scale

The primary participant fixed-effect estimate was evaluated alongside participant-day fixed effects, a broader wearable-step cohort and a strictly filtered isolated-meal sensitivity cohort. Exposure-scale analyses translate the standardized log-step coefficient into raw-step and doubling scales.

**Supplementary Table 3** | Robustness of the main post-meal step association across related analysis sets and fixed-effect specifications.

| analysis | meals | participants | estimate (95% CI) | result | scale |
|---|---|---|---|---|---|
| All wearable-step valid meals | 64,869 | 1,875 | -52.2 (-62.8, -41.5) | Estimated | mg/dL*min per 1 SD higher log post-meal steps |
| Primary analysis cohort | 55,949 | 1,627 | -53.0 (-64.2, -41.7) | Estimated | mg/dL*min per 1 SD higher log post-meal steps |
| Primary analysis, participant-day FE | 55,949 | 1,627 | -56.6 (-70.2, -43.1) | Estimated | mg/dL*min per 1 SD higher log post-meal steps |
| Strictly filtered isolated meals | 30,108 | 1,614 | -55.4 (-70.4, -40.4) | Estimated | mg/dL*min per 1 SD higher log post-meal steps |

**Supplementary Table 4** | Exposure-scale translation of the primary post-meal step association.

| model | exposure scale | meals | participants | median steps | Q1 steps | Q3 steps | estimate (95% CI) |
|---|---|---|---|---|---|---|---|
| Primary standardized log-step model | per 1 SD log1p(steps) | 55,949 | 1,627 | 354.7 | 110.0 | 851.0 | -53.0 (-64.2, -41.7) |
| Post-meal step doubling model | per doubling of 1+steps | 55,949 | 1,627 | 354.7 | 110.0 | 851.0 | -24.4 (-29.6, -19.2) |
| Raw steps per 1,000-step model | per 1000 observed steps | 55,949 | 1,627 | 354.7 | 110.0 | 851.0 | -46.1 (-56.7, -35.5) |

#### Supplementary Note 3: Dose, Timing, Meal Context and Negative Controls

Observed step-bin analyses translate the continuous model scale to practical post-meal step totals, and spline landmarks provide a smoothness check. Timing summaries use StepCount event overlap to describe where tracked 0-120 minute steps fall, but the event granularity is coarse and should not be interpreted as minute-level bout timing. Matched-meal analyses pair high-step and low-step meals within participant after balancing measured meal and timing context.

**Supplementary Table 5** | Dose-response across observed post-meal step bins, using 0-50 steps as the reference.

| post-meal step bin | meals | participants | median steps | estimate (95% CI) | result |
|---|---|---|---|---|---|
| 0-50 | 8,349 | 1,425 | 24.0 | Reference | Reference |
| 51-150 | 8,648 | 1,493 | 93.0 | 4.0 (-29.2, 37.2) | Estimated |
| 151-300 | 8,497 | 1,512 | 220.0 | -8.9 (-44.2, 26.4) | Estimated |
| 301-500 | 7,890 | 1,508 | 389.1 | -23.1 (-59.0, 12.9) | Estimated |
| 501-750 | 6,550 | 1,451 | 610.7 | -73.6 (-112.5, -34.8) | Estimated |
| 751-1,000 | 4,401 | 1,347 | 860.4 | -90.6 (-132.1, -49.1) | Estimated |
| 1,001-1,500 | 4,917 | 1,310 | 1,208.4 | -97.5 (-138.5, -56.4) | Estimated |
| 1,501-2,500 | 3,807 | 1,221 | 1,851 | -154.4 (-197.1, -111.8) | Estimated |
| 2,500+ | 2,873 | 998 | 3,570.2 | -223.9 (-276.4, -171.4) | Estimated |

**Supplementary Table 6** | Spline-based dose-response landmarks relative to 50 post-meal steps.

| post-meal steps | reference steps | meals | participants | estimate (95% CI) | scale |
|---|---|---|---|---|---|
| 50.0 | 50.0 | 55,932 | 1,610 | 0.0 (0.0, 0.0) | adjusted PPGR iAUC contrast versus 50 post-meal steps |
| 100.0 | 50.0 | 55,932 | 1,610 | -5.2 (-13.0, 2.5) | adjusted PPGR iAUC contrast versus 50 post-meal steps |
| 250.0 | 50.0 | 55,932 | 1,610 | -29.4 (-43.7, -15.1) | adjusted PPGR iAUC contrast versus 50 post-meal steps |
| 500.0 | 50.0 | 55,932 | 1,610 | -62.9 (-82.5, -43.3) | adjusted PPGR iAUC contrast versus 50 post-meal steps |
| 750.0 | 50.0 | 55,932 | 1,610 | -89.1 (-113.4, -64.8) | adjusted PPGR iAUC contrast versus 50 post-meal steps |
| 1,000 | 50.0 | 55,932 | 1,610 | -109.9 (-137.4, -82.5) | adjusted PPGR iAUC contrast versus 50 post-meal steps |
| 1,500 | 50.0 | 55,932 | 1,610 | -141.5 (-171.3, -111.8) | adjusted PPGR iAUC contrast versus 50 post-meal steps |
| 2,500 | 50.0 | 55,932 | 1,610 | -183.9 (-216.2, -151.6) | adjusted PPGR iAUC contrast versus 50 post-meal steps |
| 5,000 | 50.0 | 55,932 | 1,610 | -244.8 (-297.8, -191.8) | adjusted PPGR iAUC contrast versus 50 post-meal steps |

**Supplementary Table 7** | Distribution of tracked steps across the four 30-minute intervals after meal start.

| window (min) | meals | participants | median window steps | Q1 window steps | Q3 window steps | share of 0-120 min steps (%) | median StepCount event duration (min) | correlation with prorated total |
|---|---|---|---|---|---|---|---|---|
| 0-30 | 55,949 | 1,627 | 70.5 | 17.7 | 174.3 | 19.4 | 60.0 | 0.854 |
| 30-60 | 55,949 | 1,627 | 67.9 | 14.5 | 189.2 | 23.0 | 60.0 | 0.854 |
| 60-90 | 55,949 | 1,627 | 69.1 | 13.0 | 209.9 | 26.4 | 60.0 | 0.854 |
| 90-120 | 55,949 | 1,627 | 69.1 | 11.9 | 228.6 | 31.3 | 60.0 | 0.854 |

**Supplementary Table 8** | Carbohydrate-threshold sensitivity analyses for the primary association.

| analysis | meals | participants | estimate (95% CI) | result | scale |
|---|---|---|---|---|---|
| Primary analysis, all carbohydrate values | 55,949 | 1,627 | -53.0 (-64.2, -41.7) | Estimated | mg/dL*min per 1 SD higher log post-meal steps |
| Primary analysis, carbohydrate >=5 g | 55,949 | 1,627 | -53.0 (-64.2, -41.7) | Estimated | mg/dL*min per 1 SD higher log post-meal steps |
| Primary analysis, carbohydrate >=10 g | 53,671 | 1,627 | -56.9 (-68.5, -45.3) | Estimated | mg/dL*min per 1 SD higher log post-meal steps |
| Primary analysis, carbohydrate 5-200 g | 55,784 | 1,627 | -52.8 (-64.0, -41.5) | Estimated | mg/dL*min per 1 SD higher log post-meal steps |
| Primary analysis, carbohydrate 10-150 g | 52,607 | 1,627 | -56.6 (-68.1, -45.1) | Estimated | mg/dL*min per 1 SD higher log post-meal steps |

**Supplementary Table 9** | Meal-composition and pre-meal-state modifiers of the post-meal step association.

| analysis | meals | participants | estimate (95% CI) | result | scale |
|---|---|---|---|---|---|
| Carbohydrate grams | 55,949 | 1,627 | -10.0 (-22.5, 2.5) | Estimated | interaction: change in post-meal step association per 1 SD higher context feature |
| Carbohydrate energy fraction | 55,949 | 1,627 | -16.2 (-25.7, -6.7) | Estimated | interaction: change in post-meal step association per 1 SD higher context feature |
| Meal energy | 55,949 | 1,627 | 3.1 (-7.5, 13.6) | Estimated | interaction: change in post-meal step association per 1 SD higher context feature |
| Protein grams | 55,949 | 1,627 | 11.8 (1.6, 22.0) | Estimated | interaction: change in post-meal step association per 1 SD higher context feature |
| Fat grams | 55,949 | 1,627 | 6.4 (-2.8, 15.6) | Estimated | interaction: change in post-meal step association per 1 SD higher context feature |
| Fiber grams | 55,949 | 1,627 | 0.5 (-10.4, 11.4) | Estimated | interaction: change in post-meal step association per 1 SD higher context feature |
| Pre-meal glucose | 55,949 | 1,627 | 3.9 (-7.3, 15.1) | Estimated | interaction: change in post-meal step association per 1 SD higher context feature |
| Previous meal gap | 55,949 | 1,627 | -3.6 (-13.3, 6.1) | Estimated | interaction: change in post-meal step association per 1 SD higher context feature |

**Supplementary Table 10** | Secondary CGM outcome and curve-shape readouts for the post-meal step association.

| analysis | meals | participants | estimate (95% CI) | result | scale |
|---|---|---|---|---|---|
| Early PPGR iAUC 0-60 min / Primary analysis | 55,949 | 1,627 | -19.4 (-25.3, -13.4) | Estimated | outcome units per 1 SD higher log post-meal steps |
| Early PPGR iAUC 0-60 min / High predicted PPGR tertile | 17,429 | 1,527 | -16.1 (-27.5, -4.7) | Estimated | outcome units per 1 SD higher log post-meal steps |
| Mid PPGR iAUC 60-120 min / Primary analysis | 55,949 | 1,627 | -25.8 (-31.7, -20.0) | Estimated | outcome units per 1 SD higher log post-meal steps |
| Mid PPGR iAUC 60-120 min / High predicted PPGR tertile | 17,429 | 1,527 | -31.6 (-43.9, -19.4) | Estimated | outcome units per 1 SD higher log post-meal steps |
| Late PPGR iAUC 120-180 min / Primary analysis | 55,949 | 1,627 | -7.3 (-12.2, -2.4) | Estimated | outcome units per 1 SD higher log post-meal steps |
| Late PPGR iAUC 120-180 min / High predicted PPGR tertile | 17,429 | 1,527 | -12.2 (-21.5, -2.9) | Estimated | outcome units per 1 SD higher log post-meal steps |
| Signed PPGR AUC 0-120 min / Primary analysis | 55,949 | 1,627 | -73.0 (-87.1, -58.8) | Estimated | outcome units per 1 SD higher log post-meal steps |
| Signed PPGR AUC 0-120 min / High predicted PPGR tertile | 17,429 | 1,527 | -66.1 (-91.5, -40.7) | Estimated | outcome units per 1 SD higher log post-meal steps |
| Peak glucose rise 0-120 min / Primary analysis | 55,949 | 1,627 | -0.5 (-0.7, -0.3) | Estimated | outcome units per 1 SD higher log post-meal steps |
| Peak glucose rise 0-120 min / High predicted PPGR tertile | 17,429 | 1,527 | -0.3 (-0.6, 0.0) | Estimated | outcome units per 1 SD higher log post-meal steps |
| Time to peak 0-120 min / Primary analysis | 38,737 | 1,619 | -0.1 (-0.4, 0.3) | Estimated | outcome units per 1 SD higher log post-meal steps |
| Time to peak 0-120 min / High predicted PPGR tertile | 13,040 | 1,494 | -0.1 (-0.7, 0.5) | Estimated | outcome units per 1 SD higher log post-meal steps |
| Mean glucose change 0-120 min / Primary analysis | 55,949 | 1,627 | -0.6 (-0.8, -0.5) | Estimated | outcome units per 1 SD higher log post-meal steps |
| Mean glucose change 0-120 min / High predicted PPGR tertile | 17,429 | 1,527 | -0.6 (-0.8, -0.3) | Estimated | outcome units per 1 SD higher log post-meal steps |
| Mean glucose change 120-180 min / Primary analysis | 55,864 | 1,627 | -0.3 (-0.4, -0.1) | Estimated | outcome units per 1 SD higher log post-meal steps |
| Mean glucose change 120-180 min / High predicted PPGR tertile | 17,408 | 1,527 | -0.3 (-0.5, -0.1) | Estimated | outcome units per 1 SD higher log post-meal steps |
| Recovery slope 60-180 min / Primary analysis | 55,949 | 1,627 | 0.0 (0.0, 0.0) | Estimated | outcome units per 1 SD higher log post-meal steps |

| analysis | meals | participants | estimate (95% CI) | result | scale |
|---|---|---|---|---|---|
| Recovery slope 60-180 min / High predicted PPGR tertile | 17,429 | 1,527 | 0.0 (0.0, 0.0) | Estimated | outcome units per 1 SD higher log post-meal steps |

**Supplementary Table 11** | Matched high-step versus low-step meal contrast after within-participant matching.

| analysis | pairs | participants | median high-step meals | median low-step meals | same weekend category (%) | median hour gap | adjusted high-low PPGR (95% CI) |
|---|---|---|---|---|---|---|---|
| Within-participant matched high-step versus low-step meals | 6,919 | 1,454 | 1,290 | 100.0 | 100.0 | 1.000 | -118.7 (-150.6, -84.8) |

**Supplementary Table 12** | Covariate balance before and after matched-meal pairing.

| feature | before matching SMD | after matching SMD | before matching absolute SMD | after matching absolute SMD |
|---|---|---|---|---|
| Hour cosine | -0.475 | -0.053 | 0.475 | 0.053 |
| Hour sine | 0.397 | -0.004 | 0.397 | 0.004 |
| Prior meal gap | 0.150 | 0.031 | 0.150 | 0.031 |
| Fiber | -0.120 | -0.083 | 0.120 | 0.083 |
| Fat | -0.105 | -0.049 | 0.105 | 0.049 |
| Protein | -0.083 | -0.055 | 0.083 | 0.055 |
| Calories | -0.078 | -0.031 | 0.078 | 0.031 |
| Weekday sine | 0.054 | -0.014 | 0.054 | 0.014 |
| Weekday cosine | -0.029 | 0.005 | 0.029 | 0.005 |
| Carbohydrate | 0.020 | -0.003 | 0.020 | 0.003 |
| Prior gap missing | -0.012 | 0.000 | 0.012 | 0.000 |
| Pre-meal glucose | -0.001 | 0.004 | 0.001 | 0.004 |

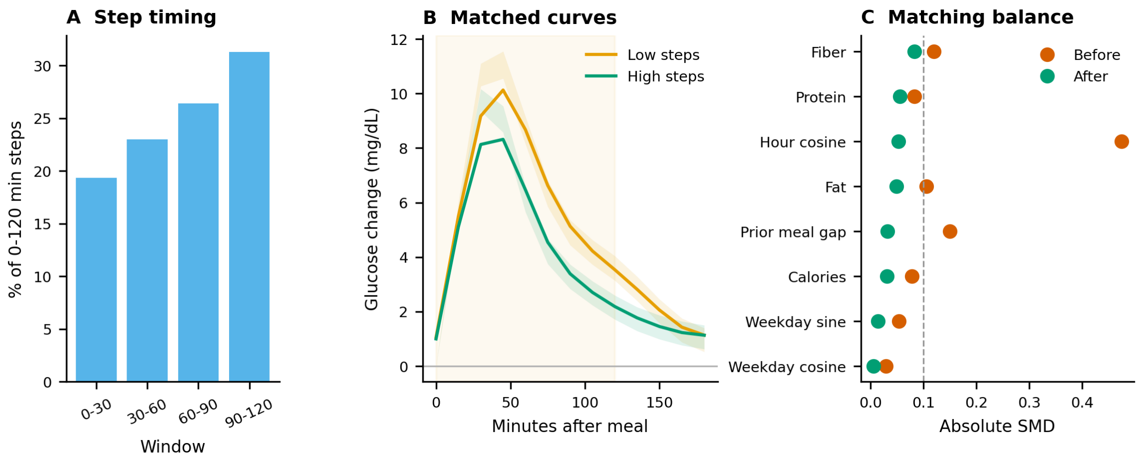


**Supplementary Figure 2** | Step timing and matched-meal checks. The timing panel reports the share of 0-120 minute steps occurring in each 30-minute window. The matched-curve panel shows meal-start baseline-subtracted glucose curves for matched high-step and low-step meals. The balance panel shows standardized mean differences before and after matching.

**Supplementary Table 13** | Negative-control outcomes for behavioral patterning and secular drift.

| analysis | meals | participants | estimate (95% CI) | scale |
|---|---|---|---|---|
| Meal protein content | 55,949 | 1,627 | -0.0 (-0.0, -0.0) | outcome units per 1 SD higher log post-meal steps |

| analysis | meals | participants | estimate (95% CI) | scale |
|---|---|---|---|---|
| Weekend meal indicator | 55,949 | 1,627 | -0.0 (-0.0, -0.0) | outcome units per 1 SD higher log post-meal steps |
| Calendar date | 55,949 | 1,627 | 0.0 (-0.0, 0.0) | outcome units per 1 SD higher log post-meal steps |

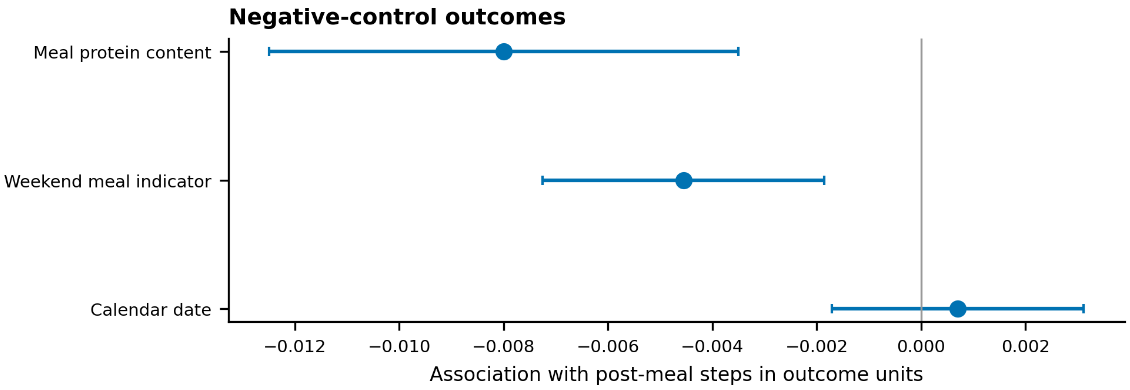


**Supplementary Figure 3** | Negative-control outcomes. Pre-exposure associations help distinguish behavioral patterning and secular drift from acute post-meal glucose effects.

#### Supplementary Note 4: Participant Context and Heterogeneity

Participant context analyses tested whether the post-meal step association varied by baseline glycemic burden, DXA adiposity and their equal-weight combination. Component-level analyses support the participant-context interpretation and are not separate primary endpoints.

**Supplementary Table 14** | Coverage of participant phenotype features used in modifier analyses.

| domain | feature | label | unit | primary analysis participants | participants with feature | coverage fraction | mean | s.d. |
|---|---|---|---|---|---|---|---|---|
| anthropometry | age_yrs | Age | years | 1,627 | 1,625 | 0.999 | 50.1 | 9.291 |
| anthropometry | bmi | BMI | kg/m2 | 1,627 | 1,622 | 0.997 | 25.2 | 3.788 |
| anthropometry | waist circumference | Waist circumference | cm | 1,627 | 1,624 | 0.998 | 85.8 | 11.8 |
| body_composition | dxa android gynoid ratio | DXA android/gynoid fat ratio | ratio | 1,627 | 1,564 | 0.961 | 0.490 | 0.191 |
| body_composition | dxa_percent_fat | DXA percent fat | % | 1,627 | 1,564 | 0.961 | 31.7 | 7.840 |
| body_composition | dxa_vat_mass | DXA visceral adipose tissue mass | g | 1,627 | 1,561 | 0.959 | 758.2 | 605.0 |

**Supplementary Table 15** | Participant-level modifiers of the post-meal step association.

| analysis | meals | participants | estimate (95% CI) | result | scale |
|---|---|---|---|---|---|
| Glycemic burden | 55,949 | 1,627 | -12.0 (-23.1, -0.8) | Estimated | interaction: change in post-meal step association per 1 SD higher modifier |
| DXA adiposity | 54,331 | 1,564 | -11.9 (-24.2, 0.4) | Estimated | interaction: change in post-meal step association per 1 SD higher modifier |
| Glycemic-adiposity | 54,331 | 1,564 | -15.6 (-27.5, -3.7) | Estimated | interaction: change in post-meal step association per 1 SD higher modifier |
| BMI | 55,742 | 1,622 | 0.6 (-11.0, 12.2) | Estimated | interaction: change in post-meal step association per 1 SD higher modifier |

| analysis | meals | participants | estimate (95% CI) | result | scale |
|---|---|---|---|---|---|
| Waist circumference | 55,838 | 1,624 | -2.6 (-14.2, 9.0) | Estimated | interaction: change in post-meal step association per 1 SD higher modifier |
| DXA android/gynoid ratio | 54,331 | 1,564 | -4.4 (-16.8, 8.0) | Estimated | interaction: change in post-meal step association per 1 SD higher modifier |
| DXA percent fat | 54,331 | 1,564 | -15.2 (-26.8, -3.5) | Estimated | interaction: change in post-meal step association per 1 SD higher modifier |
| DXA VAT mass | 54,245 | 1,561 | -6.5 (-19.6, 6.7) | Estimated | interaction: change in post-meal step association per 1 SD higher modifier |
| Sleep disturbance burden | 52,462 | 1,508 | 0.3 (-11.0, 11.6) | Estimated | interaction: change in post-meal step association per 1 SD higher modifier |
| Sleep AHI | 52,462 | 1,508 | -1.2 (-13.1, 10.7) | Estimated | interaction: change in post-meal step association per 1 SD higher modifier |
| Sleep efficiency | 52,462 | 1,508 | -4.5 (-15.6, 6.5) | Estimated | interaction: change in post-meal step association per 1 SD higher modifier |
| Median post-meal steps | 55,949 | 1,627 | -29.7 (-51.4, -8.0) | Estimated | interaction: change in post-meal step association per 1 SD higher modifier |

**Supplementary Table 16** | Post-meal step associations across glycemic-adiposity tertiles.

| analysis | meals | participants | estimate (95% CI) | result | scale |
|---|---|---|---|---|---|
| Low glycemic-adiposity | 18,502 | 522 | -32.3 (-50.1, -14.5) | Estimated | mg/dL*min per 1 SD higher log post-meal steps |
| Middle glycemic-adiposity | 18,110 | 520 | -56.4 (-77.0, -35.8) | Estimated | mg/dL*min per 1 SD higher log post-meal steps |
| High glycemic-adiposity | 17,719 | 522 | -69.0 (-90.3, -47.8) | Estimated | mg/dL*min per 1 SD higher log post-meal steps |

**Supplementary Table 17** | Participant-level heterogeneity checks for PPGR and post-meal step associations.

| analysis | meals | participants | result | interpretation |
|---|---|---|---|---|
| Observed PPGR iAUC | 55,949 | 1,627 | ICC 0.120 | Variance component summary |
| Residual PPGR after pooled step adjustment | 55,949 | 1,627 | ICC 0.199 | Variance component summary |
| Random-intercept and random step-slope model | 55,949 | 1,627 | fixed step effect -52.2; random step variance 12461.1 | Mixed-model summary |
| Mixed-model variance explained | 55,949 | 1,627 | marginal R2 0.261; conditional R2 0.477 | Mixed-model R2 summary |
| Participant-specific unadjusted within-person slopes | | 1,516 | median slope -24.4; negative share 0.550 | Participant-slope distribution |

**Supplementary Table 18** | Repeated-meal diet variability within the step-linked analysis cohort.

| label | unit | N meals / participants | n participants with ge5 meals | meal median [IQR] | participant median [IQR] | within-participant IQR median [IQR] |
|---|---|---|---|---|---|---|
| Meal energy | kcal | 55,949 / 1,627 | 1,589 | 343.7 [198.2, 566.8] | 349.6 [286.6, 429.8] | 321.8 [245.5, 424.0] |
| Carbohydrate | g | 55,949 / 1,627 | 1,589 | 36.4 [23.7, 59.3] | 37.0 [30.7, 45.6] | 31.9 [23.9, 41.2] |
| Protein | g | 55,949 / 1,627 | 1,589 | 12.4 [5.1, 25.7] | 12.7 [9.8, 16.8] | 18.4 [13.3, 24.6] |
| Fat | g | 55,949 / 1,627 | 1,589 | 12.8 [6.1, 24.3] | 13.0 [10.0, 17.0] | 16.3 [12.1, 21.3] |
| Fiber | g | 55,949 / 1,627 | 1,589 | 4.1 [1.8, 7.7] | 4.1 [3.2, 5.5] | 5.1 [3.8, 6.7] |
| Food-row count | rows | 55,949 / 1,627 | 1,589 | 3.0 [2.0, 5.0] | 3.0 [3.0, 4.0] | 2.2 [2.0, 3.0] |

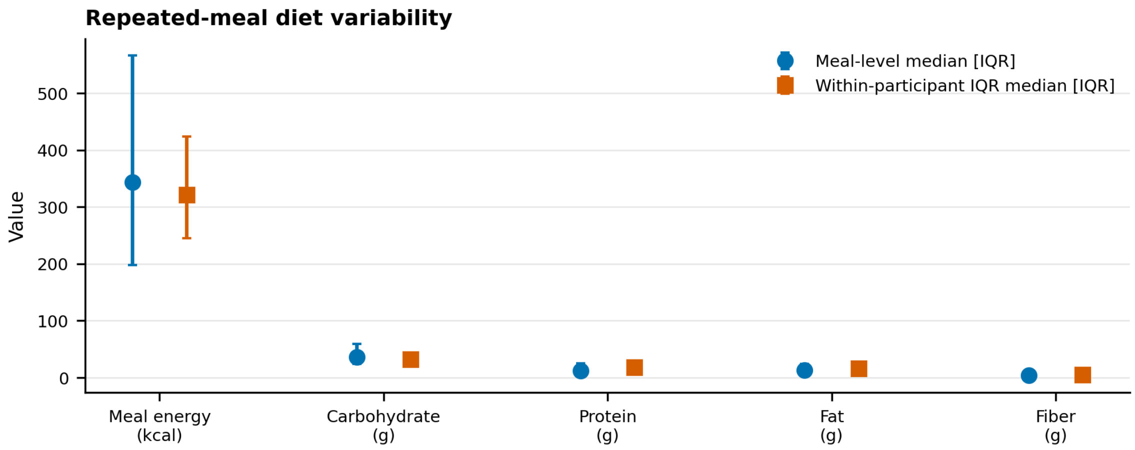


**Supplementary Figure 4** | Repeated-meal diet variability. Meal-level and within-participant summaries show dietary variation across repeated meals while reporting only aggregate statistics.

#### Supplementary Note 5: Pre-Activity Prediction and Priority Models

The pre-activity prediction model used meal, pre-meal and participant features only, excluding post-meal activity and response-derived fields. Step-response ranking analyses then tested whether pre-activity scores ordered meals by the magnitude of the observed post-meal step association.

**Supplementary Table 19** | Out-of-fold pre-activity predicted-PPGR model performance and feature-leakage check.

| analysis | meals | participants | result | check result |
|---|---|---|---|---|
| Overall out-of-fold performance | 391,214 | 9,561 | R2 0.158; r 0.398; MAE 862.9 | Estimated |
| Feature-leakage check | | | 19 of 19 features passed | Passed |

**Supplementary Table 20** | Prediction performance and participant characteristics in the broad HPP and step-linked subsets.

| analysis | meals | participants | result | interpretation |
|---|---|---|---|---|
| Broad PPGR-valid HPP frame | 391,214 | 9,561 | R2 0.158; r 0.398; MAE 862.9 | Participant-grouped out-of-fold performance for pre-activity predicted PPGR. |
| Step-linked primary analysis cohort | 55,949 | 1,627 | R2 0.163; r 0.404; MAE 835.4 | Participant-grouped out-of-fold performance for pre-activity predicted PPGR. |
| Broad PPGR-valid frame excluding step-linked meals | 335,265 | 9,289 | R2 0.157; r 0.397; MAE 867.5 | Participant-grouped out-of-fold performance for pre-activity predicted PPGR. |
| Step-linked participants - Age | | 1,627 | mean 50.05; median 49.47 | Participant characteristics for comparing the step-linked subset with the broader prediction cohort. |
| Step-linked participants - BMI | | 1,627 | mean 25.21; median 24.78 | Participant characteristics for comparing the step-linked subset with the broader prediction cohort. |
| Step-linked participants - HbA1c | | 1,627 | mean 5.32; median 5.30 | Participant characteristics for comparing the step-linked subset with the broader prediction cohort. |
| Step-linked participants - Fasting glucose | | 1,627 | mean 91.02; median 90.60 | Participant characteristics for comparing the step-linked subset with the broader prediction cohort. |

| analysis | meals | participants | result | interpretation |
|---|---|---|---|---|
| Broad PPGR-valid participants without step-linked meals - Age | | 7,934 | mean 50.90; median 50.08 | Participant characteristics for comparing the step-linked subset with the broader prediction cohort. |
| Broad PPGR-valid participants without step-linked meals - BMI | | 7,934 | mean 25.82; median 25.34 | Participant characteristics for comparing the step-linked subset with the broader prediction cohort. |
| Broad PPGR-valid participants without step-linked meals - HbA1c | | 7,934 | mean 5.34; median 5.40 | Participant characteristics for comparing the step-linked subset with the broader prediction cohort. |
| Broad PPGR-valid participants without step-linked meals - Fasting glucose | | 7,934 | mean 91.61; median 91.00 | Participant characteristics for comparing the step-linked subset with the broader prediction cohort. |

**Supplementary Table 21** | Predicted-response interaction and tertile-specific post-meal step associations.

| analysis | meals | participants | estimate (95% CI) | result | scale |
|---|---|---|---|---|---|
| Logged meal-time primary | 55,949 | 1,627 | -12.4 (-24.5, -0.3) | Estimated | change in step association per 1 SD higher predicted PPGR |
| Logged meal-time participant-day FE | 55,949 | 1,627 | -9.3 (-23.2, 4.6) | Estimated | change in step association per 1 SD higher predicted PPGR |
| Low predicted PPGR risk | 19,766 | 1,549 | -37.5 (-51.5, -23.5) | Estimated | mg/dL*min per 1 SD higher log post-meal steps |
| Middle predicted PPGR risk | 18,754 | 1,597 | -62.9 (-79.4, -46.3) | Estimated | mg/dL*min per 1 SD higher log post-meal steps |
| High predicted PPGR risk | 17,429 | 1,527 | -62.9 (-85.8, -39.9) | Estimated | mg/dL*min per 1 SD higher log post-meal steps |

**Supplementary Table 22** | Joint glycemic-adiposity and predicted-response strata.

| analysis | meals | participants | estimate (95% CI) | result | scale |
|---|---|---|---|---|---|
| Low / Low | 5,428 | 487 | -20.9 (-43.3, 1.5) | Estimated | mg/dL*min per 1 SD higher log post-meal steps |
| Low / Middle | 6,365 | 511 | -32.2 (-55.8, -8.7) | Estimated | mg/dL*min per 1 SD higher log post-meal steps |
| Low / High | 6,709 | 492 | -39.1 (-72.6, -5.6) | Estimated | mg/dL*min per 1 SD higher log post-meal steps |
| Middle / Low | 6,051 | 500 | -39.1 (-63.5, -14.7) | Estimated | mg/dL*min per 1 SD higher log post-meal steps |
| Middle / Middle | 6,334 | 516 | -73.5 (-102.7, -44.2) | Estimated | mg/dL*min per 1 SD higher log post-meal steps |
| Middle / High | 5,725 | 499 | -72.1 (-114.0, -30.3) | Estimated | mg/dL*min per 1 SD higher log post-meal steps |
| High / Low | 7,757 | 507 | -49.2 (-74.7, -23.7) | Estimated | mg/dL*min per 1 SD higher log post-meal steps |
| High / Middle | 5,504 | 514 | -82.9 (-117.6, -48.1) | Estimated | mg/dL*min per 1 SD higher log post-meal steps |
| High / High | 4,458 | 483 | -85.7 (-136.4, -35.0) | Estimated | mg/dL*min per 1 SD higher log post-meal steps |

**Supplementary Table 23** | Comparison of candidate pre-activity step-response scores.

| score | score label | bottom quintile (95% CI) | top quintile (95% CI) | top-bottom difference | step-response score interaction (95% CI) | selected for main analysis |
|---|---|---|---|---|---|---|
| Ridge residualized step-response score | Ridge step-response score | -32.1 (-49.5, -14.6) | -92.2 (-124.0, -60.4) | -60.1 | -27.3 (-38.6, -16.0) | No |
| Pre-activity predicted-response score | Pre-activity predicted-response score | -20.0 (-37.6, -2.4) | -60.4 (-91.1, -29.7) | -40.4 | -13.9 (-25.6, -2.3) | Yes |
| Person-meal step-response score | Person-meal step-response score | -15.0 (-32.2, 2.2) | -79.1 (-112.6, -45.5) | -64.1 | -24.3 (-36.8, -11.9) | Yes |

**Supplementary Table 24** | Post-meal step associations across selected step-response ranking quintiles.

| score | sensitivity analysis | step-response ranking quintile | meals | participants | Median predicted PPGR | median observed PPGR | Median post-meal steps | estimate (95% CI) |
|---|---|---|---|---|---|---|---|---|
| Person-meal step-response score | Logged meal-time primary | Q1 lowest | 10,867 | 1,138 | 848.8 | 351.0 | 356.0 | -15.0 (-32.2, 2.2) |
| Person-meal step-response score | Logged meal-time primary | Q2 | 10,866 | 1,337 | 1,115.9 | 715.5 | 351.0 | -39.3 (-58.9, -19.7) |
| Person-meal step-response score | Logged meal-time primary | Q3 | 10,866 | 1,382 | 1,233.4 | 918.0 | 367.0 | -51.3 (-74.5, -28.2) |
| Person-meal step-response score | Logged meal-time primary | Q4 | 10,866 | 1,348 | 1,387.5 | 1,167.8 | 352.0 | -76.0 (-101.6, -50.5) |
| Person-meal step-response score | Logged meal-time primary | Q5 highest | 10,866 | 1,109 | 1,647.4 | 1,660.5 | 355.0 | -79.1 (-112.6, -45.5) |
| Person-meal step-response score | Logged meal-time participant-day FE | Q1 lowest | 10,867 | 1,138 | 848.8 | 351.0 | 356.0 | -4.3 (-28.7, 20.1) |
| Person-meal step-response score | Logged meal-time participant-day FE | Q2 | 10,866 | 1,337 | 1,115.9 | 715.5 | 351.0 | -56.4 (-89.5, -23.3) |
| Person-meal step-response score | Logged meal-time participant-day FE | Q3 | 10,866 | 1,382 | 1,233.4 | 918.0 | 367.0 | -65.1 (-103.7, -26.6) |
| Person-meal step-response score | Logged meal-time participant-day FE | Q4 | 10,866 | 1,348 | 1,387.5 | 1,167.8 | 352.0 | -100.0 (-142.6, -57.3) |
| Person-meal step-response score | Logged meal-time participant-day FE | Q5 highest | 10,866 | 1,109 | 1,647.4 | 1,660.5 | 355.0 | -89.0 (-136.0, -42.0) |
| Person-meal step-response score | Logged meal-time strictly filtered isolated meals | Q1 lowest | 5,841 | 961 | 956.5 | 472.5 | 375.0 | -2.6 (-27.1, 21.9) |
| Person-meal step-response score | Logged meal-time strictly filtered isolated meals | Q2 | 5,840 | 1,218 | 1,186 | 823.5 | 351.9 | -50.0 (-80.2, -19.8) |
| Person-meal step-response score | Logged meal-time strictly filtered isolated meals | Q3 | 5,840 | 1,259 | 1,304.4 | 1,039.5 | 360.6 | -50.3 (-82.9, -17.6) |
| Person-meal step-response score | Logged meal-time strictly filtered isolated meals | Q4 | 5,840 | 1,189 | 1,448.4 | 1,269 | 356.0 | -104.4 (-138.9, -69.9) |
| Person-meal step-response score | Logged meal-time strictly filtered isolated meals | Q5 highest | 5,840 | 951 | 1,712.7 | 1,761.7 | 346.0 | -91.0 (-137.4, -44.5) |

**Supplementary Table 25** | Binary high-versus-low step contrasts within step-response ranking quintiles.

| score | step-response ranking quintile | meals | participants | low-step meals | high-step meals | estimate (95% CI) | scale |
|---|---|---|---|---|---|---|---|
| Person-meal step-response score | Q1 lowest | 7,551 | 1,044 | 4,401 | 3,150 | -68.7 (-110.7, -26.6) | PPGR iAUC difference for >=751 vs <=250 post-meal steps |
| Person-meal step-response score | Q2 | 7,550 | 1,273 | 4,428 | 3,122 | -100.7 (-149.6, -51.8) | PPGR iAUC difference for >=751 vs <=250 post-meal steps |
| Person-meal step-response score | Q3 | 7,551 | 1,338 | 4,397 | 3,154 | -113.3 (-172.8, -53.8) | PPGR iAUC difference for >=751 vs <=250 post-meal steps |
| Person-meal step-response score | Q4 | 7,550 | 1,300 | 4,479 | 3,071 | -194.8 (-255.2, -134.4) | PPGR iAUC difference for >=751 vs <=250 post-meal steps |

| score | step-response ranking quintile | meals | participants | low-step meals | high-step meals | estimate (95% CI) | scale |
|---|---|---|---|---|---|---|---|
| Person-meal step-response score | Q5 highest | 7,551 | 1,037 | 4,473 | 3,078 | -186.4 (-259.7, -113.2) | PPGR iAUC difference for >=751 vs <=250 post-meal steps |

**Supplementary Table 26** | Pre-activity feature set used for step-response scoring.

| feature | role | use in score | non-missing values | available before activity | note |
|---|---|---|---|---|---|
| Pre-activity predicted PPGR | pre-activity prediction | Used | 55,949 | Yes | Available before post-meal activity. |
| Glycemic-adiposity score | participant context | Used | 54,331 | Yes | Available before post-meal activity. |
| Glycemic burden score | participant context | Used | 55,949 | Yes | Available before post-meal activity. |
| DXA adiposity score | participant context | Used | 54,331 | Yes | Available before post-meal activity. |
| Energy | meal composition | Used | 55,949 | Yes | Available before post-meal activity. |
| Carbohydrate | meal composition | Used | 55,949 | Yes | Available before post-meal activity. |
| Protein | meal composition | Used | 55,949 | Yes | Available before post-meal activity. |
| Fat | meal composition | Used | 55,949 | Yes | Available before post-meal activity. |
| Fiber | meal composition | Used | 55,949 | Yes | Available before post-meal activity. |
| Carbohydrate fraction | meal composition | Used | 55,949 | Yes | Available before post-meal activity. |
| Logged food items per meal | meal composition | Used | 55,949 | Yes | Available before post-meal activity. |
| Previous meal gap | meal timing | Used | 55,949 | Yes | Available before post-meal activity. |
| Meal hour sine | meal timing | Used | 55,949 | Yes | Available before post-meal activity. |
| Meal hour cosine | meal timing | Used | 55,949 | Yes | Available before post-meal activity. |
| Weekday sine | meal timing | Used | 55,949 | Yes | Available before post-meal activity. |
| Weekday cosine | meal timing | Used | 55,949 | Yes | Available before post-meal activity. |
| Pre-meal baseline glucose | pre-meal glucose | Used | 55,949 | Yes | Available before post-meal activity. |
| Pre-meal glucose slope | pre-meal glucose | Unavailable | 0 | Yes | Available before post-meal activity in principle, but no non-missing values were available in the analysis feature table. |
| Age | participant characteristics | Used | 55,848 | Yes | Available before post-meal activity. |
| Female sex | participant characteristics | Used | 55,848 | Yes | Available before post-meal activity. |
| BMI | participant characteristics | Used | 55,742 | Yes | Available before post-meal activity. |
| HbA1c | participant glycemia | Used | 32,939 | Yes | Available before post-meal activity. |
| Nearest fasting glucose | participant glycemia | Used | 52,594 | Yes | Available before post-meal activity. |
| 0-120 min steps | primary exposure | Excluded | 55,949 | No | Excluded from score inputs by design. |
| Log-transformed 0-120 min steps | exposure transform | Excluded | 55,949 | No | Excluded from score inputs by design. |
| 120-240 min steps | future-activity control | Excluded | 55,949 | No | Excluded from score inputs by design. |
| Log-transformed 120-240 min steps | future-activity transform | Excluded | 55,949 | No | Excluded from score inputs by design. |

| feature | role | use in score | non-missing values | available before activity | note |
|---|---|---|---|---|---|
| PPGR iAUC 0-120 min | target outcome | Excluded | 55,949 | No | Excluded from score inputs by design. |
| High-vs-low step evaluation exposure | evaluation exposure | Excluded | 38,924 | No | Excluded from score inputs by design. |

**Supplementary Table 27** | Pre-meal step sensitivity analyses.

| label | meals | participants | post-meal estimate (95% CI) | pre-meal step estimate (95% CI) | interpretation |
|---|---|---|---|---|---|
| Primary analysis reference | 55,949 | 1,627 | -53.0 (-64.2, -41.7) | | Sensitivity adjustment; pre-meal steps can precede meal choice and post-meal behavior. |
| Primary analysis adjusted for -60 to 0 min steps | 55,949 | 1,627 | -62.1 (-73.5, -50.6) | 20.4 (15.5, 25.2) | Sensitivity adjustment; pre-meal steps can precede meal choice and post-meal behavior. |
| Primary analysis participant-day FE reference | 55,949 | 1,627 | -56.6 (-70.2, -43.1) | | Sensitivity adjustment; pre-meal steps can precede meal choice and post-meal behavior. |
| Primary analysis participant-day FE adjusted for -60 to 0 min steps | 55,949 | 1,627 | -61.0 (-74.6, -47.3) | 15.0 (9.2, 20.7) | Sensitivity adjustment; pre-meal steps can precede meal choice and post-meal behavior. |

**Supplementary Table 28** | Meal time-of-day sensitivity analyses.

| analysis | label | meal-time definition | meals | participants | estimate (95% CI) | difference from primary estimate | joint clock-time interaction P | result |
|---|---|---|---|---|---|---|---|---|
| Primary model with meal-hour adjustment | Primary model with cyclic meal-hour adjustment | all logged meal times | 55,949 | 1,627 | -53.0 (-64.2, -41.7) | 0.000 | | Estimated |
| Primary model without meal-hour adjustment | Primary model without cyclic meal-hour adjustment | all logged meal times | 55,949 | 1,627 | -48.4 (-59.4, -37.5) | 4.523 | | Estimated |
| Step x meal-hour sine interaction | Step x meal-hour sine interaction | continuous cyclic meal hour | 55,949 | 1,627 | 7.1 (-10.5, 24.6) | | 0.001 | Estimated |
| Step x meal-hour cosine interaction | Step x meal-hour cosine interaction | continuous cyclic meal hour | 55,949 | 1,627 | -26.2 (-44.6, -7.7) | | 0.001 | Estimated |
| Morning meals | Morning meals (05:00-10:59) | 05:00-10:59 | 13,348 | 1,531 | -28.7 (-45.5, -11.8) | 24.3 | | Estimated |
| Midday meals | Midday meals (11:00-15:59) | 11:00-15:59 | 19,882 | 1,599 | -42.1 (-60.4, -23.9) | 10.8 | | Estimated |
| Evening meals | Evening meals (16:00-21:59) | 16:00-21:59 | 20,211 | 1,591 | -54.8 (-71.6, -38.0) | -1.808 | | Estimated |
| Late-night meals | Late-night meals (22:00-04:59) | 22:00-04:59 | 2,508 | 965 | -42.7 (-110.0, 24.6) | 10.2 | | Estimated |

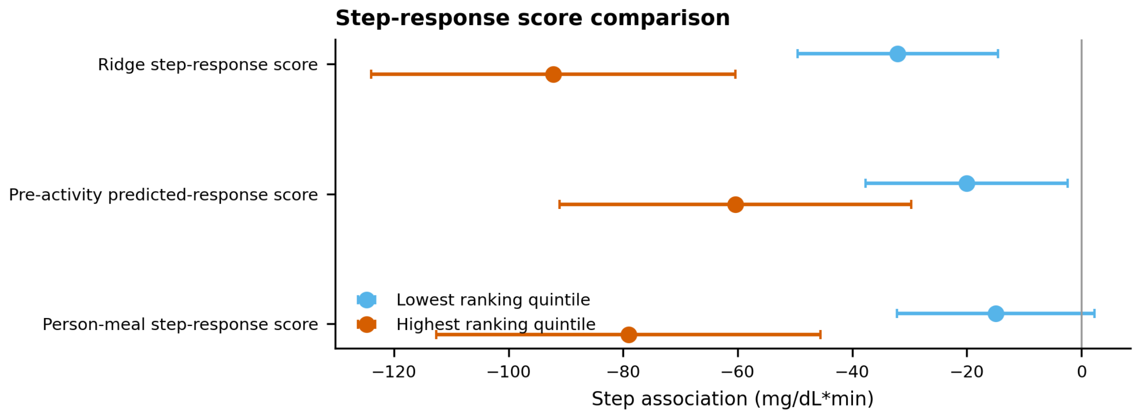


**Supplementary Figure 5** | Step-response score comparison. Points compare bottom and top ranking-quintile post-meal step associations for each candidate pre-activity score; the selected person-meal score is used in the main manuscript.
